# Longitudinal multi-omics analysis identifies responses of megakaryocytes, erythroid cells and plasmablasts as hallmarks of severe COVID-19 trajectories

**DOI:** 10.1101/2020.09.11.20187369

**Authors:** Joana P. Bernardes, Neha Mishra, Florian Tran, Thomas Bahmer, Lena Best, Johanna I. Blase, Dora Bordoni, Jeanette Franzenburg, Ulf Geisen, Jonathan Josephs-Spaulding, Philipp Köhler, Axel Künstner, Elisa Rosati, Anna C. Aschenbrenner, Petra Bacher, Nathan Baran, Teide Boysen, Burkhard Brandt, Niklas Bruse, Jonathan Dörr, Andreas Dräger, Gunnar Elke, David Ellinghaus, Julia Fischer, Michael Forster, Andre Franke, Sören Franzenburg, Norbert Frey, Anette Friedrichs, Janina Fuß, Andreas Glück, Jacob Hamm, Finn Hinrichsen, Marc P. Hoeppner, Simon Imm, Ralf Junker, Sina Kaiser, Ying H. Kan, Rainer Knoll, Christoph Lange, Georg Laue, Clemens Lier, Matthias Lindner, Georgios Marinos, Robert Markewitz, Jacob Nattermann, Rainer Noth, Peter Pickkers, Klaus F. Rabe, Alina Renz, Christoph Röcken, Jan Rupp, Annika Schaffarzyk, Alexander Scheffold, Jonas Schulte-Schrepping, Domagoj Schunck, Dirk Skowasch, Thomas Ulas, Klaus-Peter Wandinger, Michael Wittig, Johannes Zimmermann, Hauke Busch, Bimba Hoyer, Christoph Kaleta, Jan Heyckendorf, Matthijs Kox, Jan Rybniker, Stefan Schreiber, Joachim Schultze, Philip Rosenstiel, HCA Lung Biological Network and the Deutsche COVID-19 Omics Initiative (DeCOI)

## Abstract

The pandemic spread of the potentially life-threatening disease COVID-19 requires a thorough understanding of the longitudinal dynamics of host responses. Temporal resolution of cellular features associated with a severe disease trajectory will be a pre-requisite for finding disease outcome predictors. Here, we performed a longitudinal multi-omics study using a two-centre German cohort of 13 patients (from Cologne and Kiel, cohort 1). We analysed the bulk transcriptome, bulk DNA methylome, and single-cell transcriptome (>358,000 cells, including BCR profiles) of peripheral blood samples harvested from up to 5 time points. The results from single-cell and bulk transcriptome analyses were validated in two independent cohorts of COVID-19 patients from Bonn (18 patients, cohort 2) and Nijmegen (40 patients, cohort 3), respectively. We observed an increase of proliferating, activated plasmablasts in severe COVID-19, and show a distinct expression pattern related to a hyperactive cellular metabolism of these cells. We further identified a notable expansion of type I IFN-activated circulating megakaryocytes and their progenitors, indicative of emergency megakaryopoiesis, which was confirmed in cohort 2. These changes were accompanied by increased erythropoiesis in the critical phase of the disease with features of hypoxic signalling. Finally, projecting megakaryocyte- and erythroid cell-derived co-expression modules to longitudinal blood transcriptome samples from cohort 3 confirmed an association of early temporal changes of these features with fatal COVID-19 disease outcome. In sum, our longitudinal multi-omics study demonstrates distinct cellular and gene expression dynamics upon SARS-CoV-2 infection, which point to metabolic shifts of circulating immune cells, and reveals changes in megakaryocytes and increased erythropoiesis as important outcome indicators in severe COVID-19 patients.

## Introduction

Coronavirus disease 2019 (COVID-19), caused by infection with severe acute respiratory syndrome coronavirus 2 (SARS-CoV-2), is rapidly emerging into a pandemic resulting in more than 27 million cases (as of 10^th^ Sep, 2020) and a death toll of more than 900,000 people globally (https://coronavirus.jhu.edu/map.html). Since its emergence in Wuhan, China in December 2019 ^1,2^, it has spread globally by aerosol and droplet-mediated human-to-human transmission ^3-5^. COVID-19 shows a heterogeneous clinical course, ranging from asymptomatic carriers to severe cases with overwhelming inflammatory responses leading to organ failure and death.

SARS-CoV-2 infection - like other coronavirus infections - normally triggers a self-limiting protective immune response via macrophages and monocytes that respond to the infection and foster an adaptive T and B cell reaction. Severe, and ultimately fatal, COVID-19 is associated with a dysfunctional immune response, characterized by systemic inflammation and widespread inflammatory lung affection. Critically ill COVID-19 patients show higher blood plasma levels of numerous cytokines and chemokines (often termed “cytokine storm”), e.g., granulocyte colony-stimulating factor (GM-CSF), tumour necrosis factor (TNF), IL-6, CC-chemokine ligand CCL2 and CXC-chemokine ligand 10 (CXCL10) ^2,6-8^. Similar to other Betacoronaviruses, a profoundly impaired interferon (IFN) type I response with low interferon production and heterogeneous regulation of interferon-stimulated genes has been suggested to influence the course of severe COVID-19 ^9,10^. Immunophenotyping studies have demonstrated several cellular hallmarks of the systemic inflammatory response to SARS-CoV-2 infection. A significant expansion of populations of IL-6 secreting CD14^hi^CD16^hi^ monocytes as well as a decrease of non-classical (CD14^lo^CD16^hi^) monocytes was shown in the peripheral blood of severe COVID-19 patients^10-14^. While a wide upregulation of inflammatory cytokine mRNA levels was not detectable in monocytic populations by single-cell RNA sequencing (scRNA-seq) ^15^, another study demonstrated a broad impairment of the myeloid lineages ^14^. Hyperinflammatory COVID-19 disease behaviour was associated with the appearance of proliferating, type I IFN activated CD14^+^HLA^lo^ suppressive monocytes and emergency granulopoiesis with elevated pre-neutrophil counts. Another common feature in many patients with severe COVID-19 is T cell lymphopenia, which is more pronounced in the CD8^+^ T cell compartment ^2,16-19^. Several reports have highlighted the functionally exhausted phenotype of remaining T cells ^7,17,20-22^. SARS-CoV-2 infection has been shown to elicit specific T cell and B cell responses, indicated by the appearance of SARS-CoV-2 peptide-specific T cells ^23,24^ and the production of SARS-CoV-2 specific antibody responses ^25,26^, of which, however, the dynamics concerning disease outcome and lasting immunity are only incompletely understood.

A significant disease burden is mounted by thrombotic complications associated with COVID-19. Thromboembolic events, including pulmonary embolism or thrombosis, are a frequent clinical feature of critically ill COVID-19 cases ^12,19^, sometimes despite sufficient anticoagulation. The diffuse alveolar damage in SARS-CoV-2 infection has been linked to the formation of vascular thrombi in the lungs ^27^ with alveolar capillary microthrombi nine times more prevalent in patients with COVID-19 than in influenza autopsies. Endothelial injury, endothelialitis and microthrombotic complications ^28^ also affect other organ systems, e.g. heart, liver, kidney and brain ^29^. Patients exhibit elevated D-dimer levels, thrombocytopenia, complement activation, and thrombotic microvascular injury ^30^. Predisposition to this severely heightened state of coagulation might be linked to direct endothelial damage, hypoxia, immobilization, diffuse intravascular coagulation, but also might be a consequence of the hyperinflammatory state *per se*. Several studies have suggested platelet hyperreactivity, pathological-platelet immune cell interactions ^31^, and presence of megakaryocytes in affected lungs ^32^ as a determinant of severe COVID-19. However, a comprehensive view on the dynamics of thrombopoiesis and its association with disease course is missing.

The clinically heterogeneous disease presentation renders individual molecular dynamics of the haematopoietic and immune cell compartments system in response to COVID-19 an important topic to understand the pivot points of the disease.

As until now, omics-driven studies have used a cross-sectional, “single-layer” design to compare cellular and molecular states between mild and severe COVID-19, there is an urgent need for detailed insights into longitudinal trajectories of the disease in an integrated multi-omics approach and for comparing such high-density molecular data sets with detailed clinical characteristics of the individual patients. Two recent studies have demonstrated the 33,34. We power of longitudinal analysis using mostly flow cytometry-based methods hypothesized that a deep understanding of individual dynamics of functional genomic states of cellular compartments in the peripheral blood not only depicts the systemic response to SARS-CoV-2 but also may shed light on the importance of bone marrow precursor mobilization as a fundamental feature in critically ill COVID-19 patients.

## Results

To elucidate cellular patterns and activated pathways of the systemic immune response to COVID-19, we applied a multi-omics approach using up to 5 longitudinal peripheral blood samples of 13 hospitalized patients and one additional recovery control from two University hospitals in Germany (Cologne and Kiel). To identify functional changes associated with moderate to severe disease trajectories in peripheral blood mononuclear cells (PBMCs) from these individual patients, we employed parallel single-cell RNA sequencing (scRNA-seq, 10x Genomics), single-cell BCR profiling, bulk mRNA sequencing (RNA-seq), BCR amplicon sequencing and multicolour flow cytometry. DNA methylation profiling by Illumina Bead Arrays (hereafter EPIC array) and multiplex cytokine ELISA analysis from serum was performed in a subset of seven patients (Figure 1a). All patients were recruited at admission and samples were taken systematically at days 0, 2, 7, 10, 13 and/or at discharge. Three patients were diagnosed with Acute Respiratory Distress Syndrome (ARDS), two of which had a fatal disease course. Five patients received remdesivir after inclusion into this study. Patient demographics and clinical characteristics are described in Table 1 and individual detailed clinical data are reported in Supplementary Table 1. 14 age- and gender-matched healthy controls were processed in parallel. To describe the heterogenous disease trajectories over time, we used a modified WHO ordinal scale ^35^, which also considers the behaviour of several inflammatory markers (serum CRP, serum IL-6 and ferritin, Table 2). The score was used to classify patients along their disease course (Figures 1b and 1c) enabling the interrogation of molecular states associated with transition between phases (e.g., complicated to early convalescent). Phases were defined as pseudotimes in accordance with WHO and the LEOSS register (www.leoss.net) to depict the longitudinal course of the disease: incremental (pseudotime 1, where clinical symptoms and inflammatory markers were increasing, ICU or non-ICU), critical (pseudotime 2, ICU, mechanically ventilated with signs of ARDS), complicated (pseudotime 3, state with severe signs of a systemic inflammatory response, ICU, high-flow oxygen, intubation readiness), moderate or early convalescent (pseudotime 4, supplemental oxygen, significant signs of systemic inflammation), late convalescent (pseudotime 5, intermittent supplemental oxygen, minor signs of inflammation), recovery/pre-discharge (pseudotime 6, no supplemental oxygen, absent inflammation markers) and long-term follow-up (pseudotime 7, at least two weeks after hospital discharge) (Table 3). Note that pseudotimes 2 and 3 do not directly reflect the chronological order, but can also represent peak levels, i.e. a given patient might have gone from 1 to 3 without intubation/mechanical ventilation.

**Table 1.** Clinical characteristics of the patients in cohort 1 (related to Figure 1)

| Patient-ID | 001 | 002 | 003 | 004 | 005 | 006 | 007 |
| --- | --- | --- | --- | --- | --- | --- | --- |
| Sex | male | male | female | male | male | male | male |
| Ethnicity | caucasian | caucasian | caucasian | caucasian | caucasian | caucasian | caucasian |
| Age (range) | 70-79 | 50-59 | 80-89 | 60-69 | 40-49 | 70-79 | 20-29 |
| Smoker | ex smoker (50 PY) | no | no | no | no | no | no |
| Co-morbidities | HTN, COPD, Ath | - | CRC (metastatic) | HTN | A, PSO, T2DM | HTN | - |
| WHO scale (max) | 4 | 8 | 8 | 4 | 4 | 4 | 2 |
| CRP range [mg/l] | 9.0 - 100.1 | 12.96 - 358.0 | 50.6 - 169.0 | 34.3 - 241.0 | 4.3 - 168.0 | 0.5 - 49.2 | - |
| IL-6 range [ng/l] | 14.1 - 135.0 | 52.6 - 2905.0 | 289.0 - 579.0 | 41.7 - 324.0 | 4.2 - 173.0 | 17.6 - 69.0 | - |
| Ferritin range [µg/ml] | 710 - 2725 | 1295 - 8000 | 1118 - 1224 | 1239 - 2354 | 457 - 1184 | 666 - 2378 | - |
| Max disease pseudotime | early conval. | critical | critical | complicated | complicated | recovery | recovery |
| Recovered | yes | no | no | yes | yes | yes | yes |
| COVID-19-specific treatment | remdesivir | remdesivir | - | remdesivir | remdesivir | remdesivir | - |
| Anti-infective treatment | sultamicillin | pip/taz | pip/taz, cipro | sulta | none | none | none |

| Patient-ID | 008 | 009 | 010 | 011 | 012 | 013 | 014 |
| --- | --- | --- | --- | --- | --- | --- | --- |
| Sex | female | male | male | female | female | female | female |
| Ethnicity | caucasian | caucasian | caucasian | caucasian | caucasian | caucasian | asian |
| Age (range) | 60-69 | 50-59 | 70-79 | 70-79 | 50-59 | 60-69 | 50-59 |
| Smoker | no | no | no | no | no | no | no |
| Co-morbidities | post renal transplant, GN, HTN | MESH, Metabolic syndrome | Renal failure, HTN, T2DM, osteomyelitis, Ath | AF, multiple CVA | CAPA, candidemia, sepsis, AVI, Herpes labialis | Obesity, HTN, T2DM, post breast cancer | T2DM, schizophr., HIV infection |
| WHO scale (max) | 4 | 4 | 4 | 3 | 6 | 4 | 5 |
| CRP range [mg/l] | 41.9 - 217.3 | 5.5 - 115.7 | 41.7 - 287.6 | 0.5 - 6.1 | 2.1 - 347.5 | 1.0 - 144.2 | 25.8 - 110.6 |
| IL-6 range [ng/l] | 9.0 - 43.0 | 4.0 - 38.0 | 31.0 - 574.0 | 2.0 - 6.0 | 62.0 - 311.0 | 48.0 - 70.0 | 27.0 - 46.0 |
| Ferritin range [µg/ml] | 909-1071 | 1618-1901 | 802-4574 | 210 - 331 | 368 - 1090 | 815 - 2336 | 216 - 711 |
| Max disease pseudotime | complicated | early conval. | complicated | recovery | critical | complicated | complicated |
| Recovered | yes | yes | yes | yes | yes | yes | yes |
| COVID-19-specific treatment | - | - | - | - | ribavirin + lopinavir/r | tocilizumab | tocilizumab |
| Anti-infective treatment | none | none | none | none | caspo | none | none |

Abbreviations:
PY: Pack years, HTN: arterial hypertension, CAPA: COVID-19-associated pulmonary aspergillosis, COPD: Chronic obstructive pulmonary disease, Ath: Atherosclerosis, CRC: colorectal cancer, A: Asthma, PSO: psoriasis, T2DM: type II diabetes mellitus, GN: glomerulonephritis, MESH: metabolic steatohepatitis, AF: atrial fibrillation, CVA: cerebrovascular accidents, AVI: aortic valve insufficiency, schizophr.: schizophrenia, HIV: human immunodeficiency virus, conval.: convalescence, /r: ritonavir, pip/taz: piperacillin/tazobactam, caspo: caspofungin

**Figure 1.**
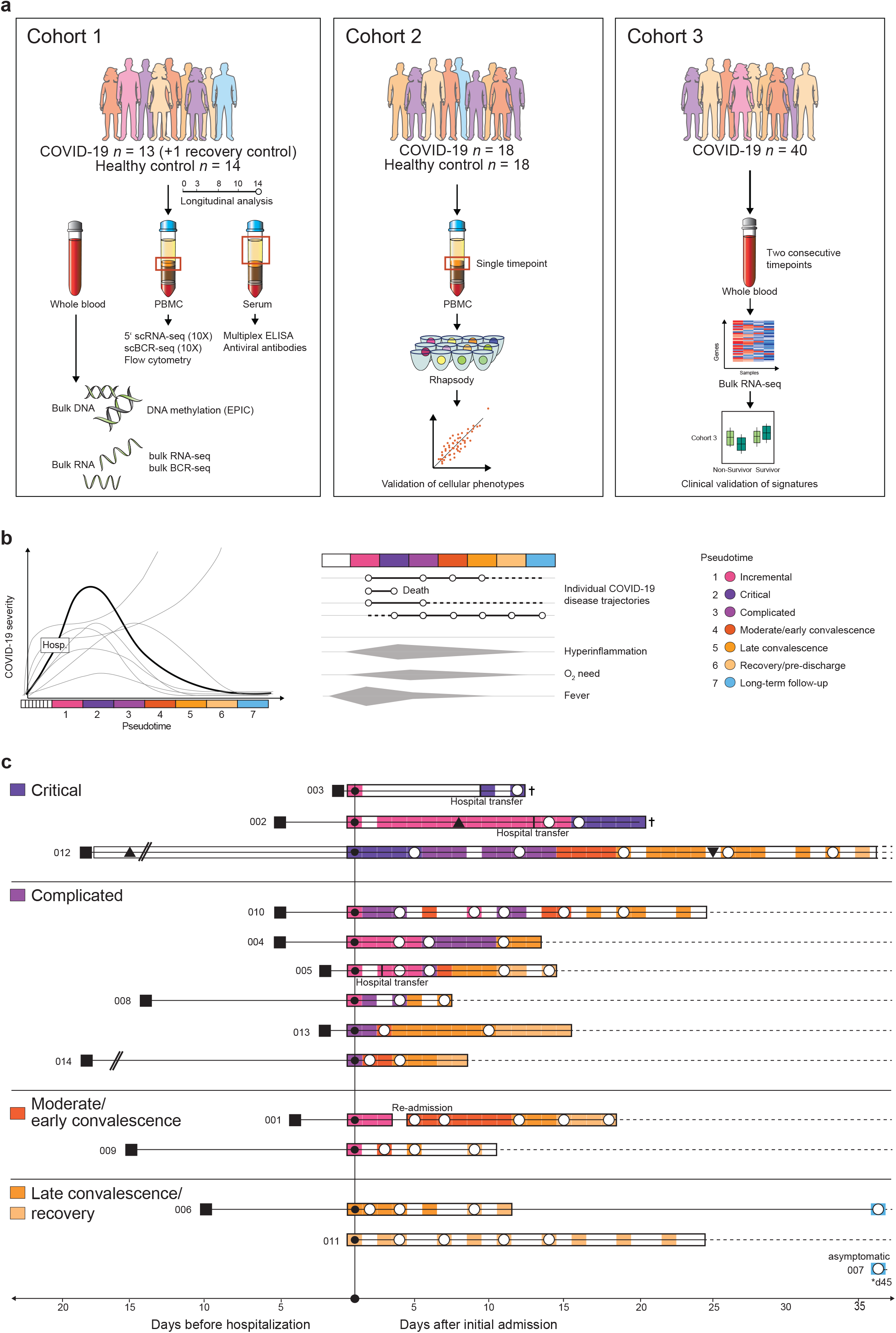
Clinical definition of disease phases for disease trajectory analyses. **a**, Overview of the cohorts used in this study. Cohort 1 underwent longitudinally multi-omics phenotyping. Cohort 2 served as an independent cross-sectional validation cohort for the cellular phenotypes observed in cohort 1. Cohort 3 was used as an independent validation cohort for gene signature-clinical outcome associations. **b**, Concept of disease phase pseudotimes. Clinical disease phases based on inflammatory markers and ventilation need (according to WHO ordinal scale) were defined. Pseudotimes reflect temporal disease severity and distinguish between incremental and recovering disease stages. Hosp. = Hospitalization. **c**, Overview of the cohort 1. All patients are temporally aligned to the day of initial admission due to COVID-19 complications. Squares mark the day of COVID-19-related symptom onset. Frames mark days of in-patient care, while the colour is representing the disease pseudotime. Sampling days were marked with a white circle. Intubations and extubations are depicted with triangle symbols, if applicable. For detailed patient characteristics see Table 1 and Supplementary Table 1.

**Figure 2.**
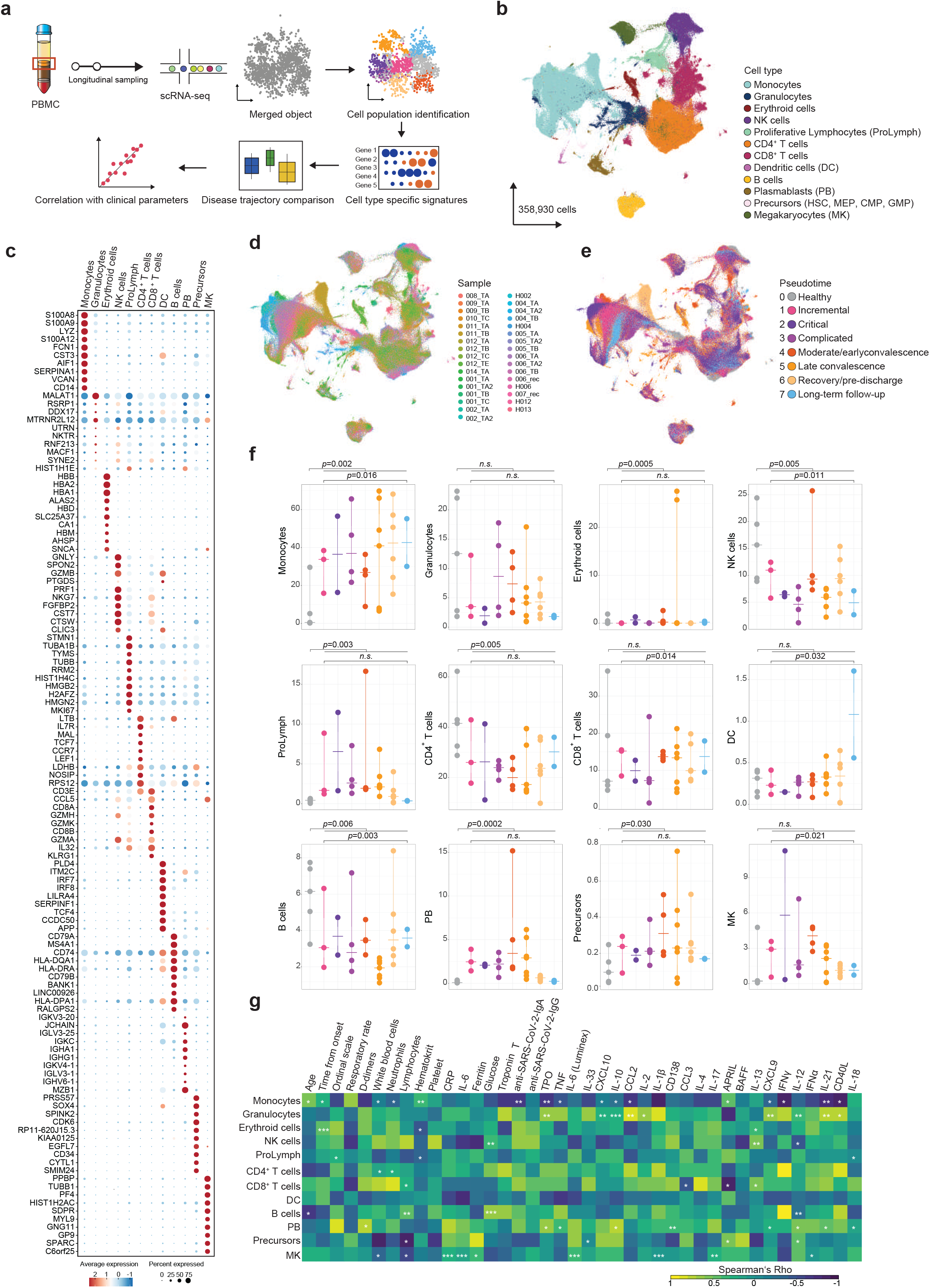
scRNA-seq analysis identifies cellular changes along COVID-19 disease trajectories. **a**, Schematic workflow for the scRNA-seq analyses performed on PBMCs. **b**, Cell type UMAP representation of all merged samples. Twelve cell types were identified by cluster gene signatures. In total, 358,930 cells are depicted. **c**, Sample of origin UMAP representation of all merged samples. Cells were coloured by the sample. Samples nomenclature based on patient ID (001-014) and time points of sample collection day 1 (after admission TA), day 3 (TA2), day 8 (TB), day 11 (TB2), day 14 (TC) and day 20 (TE). **d**, Pseudotime UMAP representation of all merged samples. Cells were coloured by pseudotime. **e**, Dot plot for cell type-specific signature genes. Genes were selected based on ten most characteristic upregulated genes. Colour discriminates upregulated (red) or downregulated (blue) genes, while point sizes represents the number of cells per group expressing the corresponding gene. **f**, Cell-specific proportions grouped by pseudotime. Points represent individual samples and horizontal bars the mean of a particular pseudotime. Pseudotimes are represented by colours. p-values based on longitudinal linear mixed model for comparison among COVID-19 pseudotimes and p-values based on Mann-Whitney test for comparisons between healthy and COVID-19 samples are reported. **g**, Correlation heatmap between cell-specific proportions and clinical parameters included in routine tests and multiplex ELISA. * indicates *p*-value < 0.05, ** *p*-value < 0.01 and *** *p*-value < 0.001 in Spearman’s correlation. Colour intensity correspond to correlation coefficient. Abbreviations: DC = dendritic cells, PB = plasmablasts, MK = megakaryocytes, HSC = hematopoietic stem cell, MEP = megakaryocyte-erythroid progenitor cell, CMP = common myeloid progenitor cell, GMP = granulocyte-monocyte progenitor.

### scRNA-seq analysis identifies cellular changes along the COVID-19 disease trajectory

We first analyzed scRNA-seq data from 358,930 cells with 10,900 cells on average per sample (Figure 2a, Supplementary Table 2). Up to four longitudinal samples were analysed per patient. Two-dimensional data representation using Uniform Manifold Approximation and Projection (UMAP) was used to visualize the structure of cellular populations (Figure 2b, 2d and 2e). Graph-based clustering identified 37 distinct cell clusters in the data set (Supplementary Figure 1a). Cell type classification was performed based on signature genes of each cluster and confirmed using reference transcriptomes available within SingleR R package ^15,36,37^ (Supplementary Figure 1b). This annotation resulted in the identification of all major cell types of the PBMC compartment (Figure 2b). The top 10 differentially expressed transcripts of each cell cluster of the UMAP are depicted in Figure 2c. Cells from individual patients were dispersed evenly in the UMAP representation, with the exception of a higher degree of interindividual heterogeneity in the monocytic subpopulations (Figure 2d). Substantial phenotypic differences could be depicted by dimensionality reduction between COVID-19 patient pseudotimes and but also between all COVID-19 patients and control samples. The changes were most overt in plasmablasts/-cells, monocytes, CD8^+^ T cells and in a cell-cluster identified as circulating megakaryocytes (Figure 2e).

Next, we quantified shifts in relative cell type proportions according to COVID-19 disease trajectories (Figure 2f). We observed a depletion of several cell types including NK cells and dendritic cells at the peak of disease severity similar to previous studies ^14,15^. Lymphopenia of both CD4^+^ and CD8^+^ T cells was present, but less pronounced in our data set during high disease activity. We also noted increased plasmablast proportions from the incremental phase to late convalescence, while B cells were depleted during early and late convalescence. Whilst the critical group (pseudotime 2) did not reach formal significance, possibly due to sample size, the data overall confirmed a strong and lasting activation of B cell immunity with an increase of circulating plasmablasts ^13,15^. Of note, we recognized the increased presence of several bone marrow-derived precursor cell types in the peripheral blood of COVID-19 patients. Prominently, megakaryocyte proportions were elevated throughout the course of the disease. To define potential cell populations, which might be directly infected by SARS-CoV2, we specifically interrogated *ACE2* mRNA levels in the data set, which were below detection limit (<7 reads) in all cell populations (data not shown).

Next, we were interested, whether changes in cell numbers correlated with clinical parameters. A correlation analysis of relative cell proportions with clinical activity parameters and a multiplex serum ELISA revealed that increased bone marrow precursor cells were associated with increased levels of IL-33, a T_h_2 cytokine involved in regulating haematopoietic stem cell regeneration ^38^ and that elevated megakaryocytes were linked to heightened inflammatory parameters, including serum CRP, IL-6 (clinical routine and multiplex ELISA), IL-1β, IL-17 and IFN-α. Plasmablast proportions were correlated with serum levels of TNF-α, IL-10, CCL3, CXCL9 and IL-21, a factor critically involved in B cell differentiation to plasmablasts via STAT3 and BLIMP-1 (PRDM1) ^39,40^ (Figure 2g).

### Whole-blood transcriptome signatures vary dynamically along COVID-19 disease trajectory

To delineate the transcriptional response to SARS-CoV-2 infection at higher temporal resolution and to instruct further cell type selection from the single-cell data set, we next analysed whole-blood transcriptomic data of the 13 hospitalized patients from up to five timepoints along with the recovery control and compared the signatures against 14 healthy controls (Figure 3a). Principal component analysis showed a separation between healthy controls and COVID-19 patients on the first principal component (PC1) (Figure 3b). Samples from COVID-19 patients taken at asymptomatic stages or after complete recovery (pseudotimes 6 and 7) also clustered with the healthy controls. Among the different samples from the same patient, critical and complicated timepoints (pseudotimes 2 and 3) tended to be located much further from the healthy controls than the milder and recovering timepoints on PC1 (Figure 3b).

**Figure 3.**
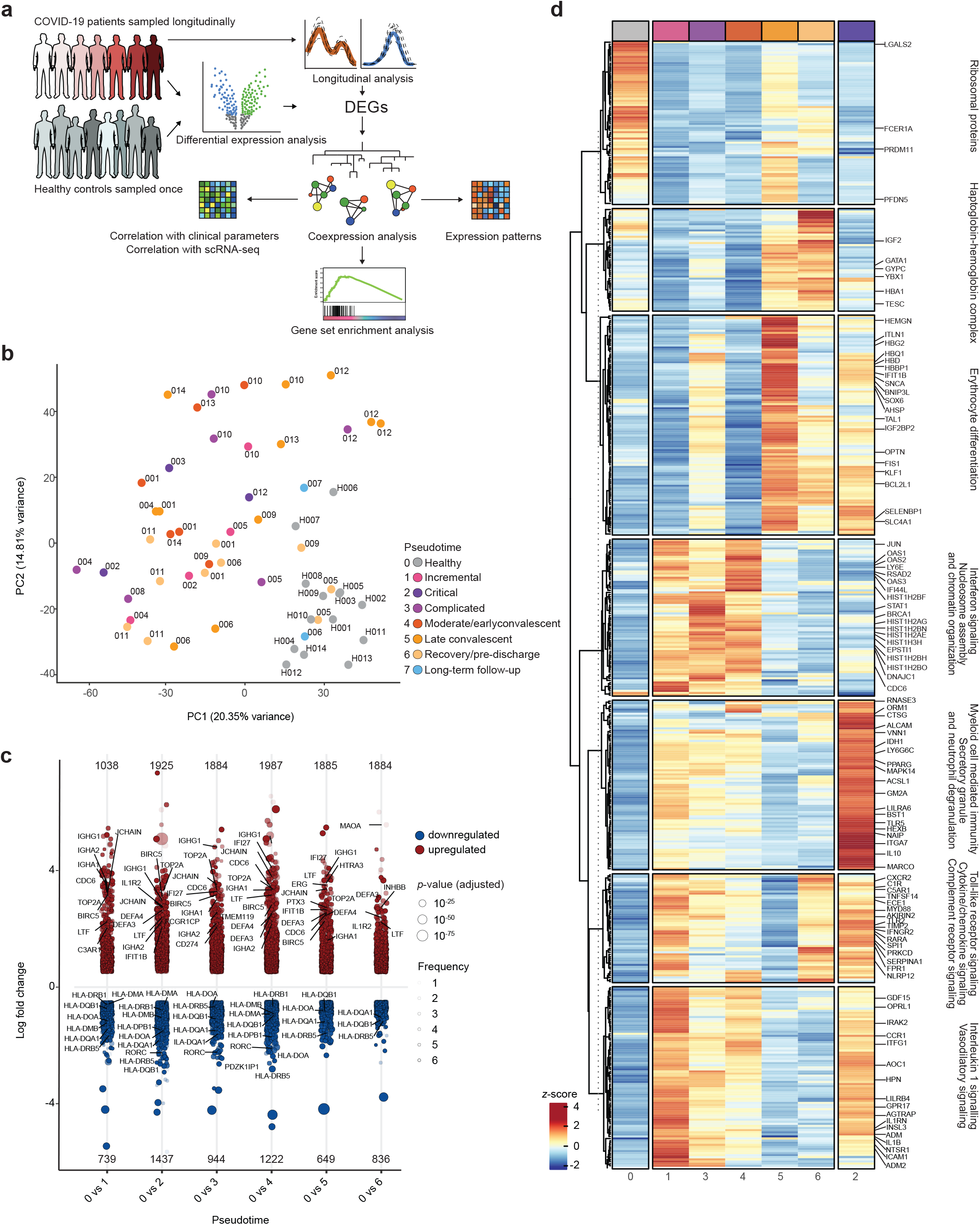
Dynamics of whole blood gene expression in COVID-19. **a**, Schematic workflow of the analysis performed on bulk RNA-seq data from whole blood. **b**, PCA plot of all control and COVID-19 samples based on the expression of all genes. Samples are colour-coded by their pseudotime and labelled by the patient or individual ID. **c**, Log_2_-fold change (y-axis) of differentially expressed genes (DEGs) between controls and each of the COVID-19 pseudotimes (x-axis). Upregulated genes are denoted in red and downregulated genes in blue. Size of the individual points is proportional to their statistical significance (adjusted *p*-value) with larger points being more significant. The transparency of the points denotes that number of comparisons in which the genes is significantly differentially expressed. The positions of individual points are jittered horizontally in order to show the density of the data. The numbers of up- and downregulated genes are written at the top and bottom of the plot, respectively. **d**, Heatmap of top 500 longitudinal DEGs across COVID-19 pseudotimes (pseudotimes 1, 3, 4, 5 and 6). Gene expression in controls and pseudotime 2 are shown for comparison on the left and right ends of the heatmap, respectively. The normalized genes counts are scaled by row and row-wise z-scores are plotted in the heatmap. Genes are hierarchically clustered using their adjacency scores as distance. (See also Supplementary Figure 2)

We next performed pairwise differential expression analyses between controls and each of the disease trajectory pseudotimes. In total, 5,915 genes were differentially expressed between healthy controls and COVID-19 patients considering all different pseudotimes (Figure 3c, Supplementary Figure 2a). Most of the differentially expressed genes (DEGs) were upregulated in COVID-19 patients and upregulated genes also had greater fold changes than downregulated genes (Figure 3c, Supplementary Figure 2a). The number of differentially expressed genes and their fold changes also varied among the different disease pseudotimes, with only 7 differentially expressed genes differentiating between healthy subjects and the long-term follow-up timepoint (pseudotime 7) and relatively smaller fold changes at late convalescence and the pre-discharge timepoint (pseudotimes 5 and 6) (Figure 3c, Supplementary Figure 2a). Notably, early changes comprised upregulation of immunoglobulin transcripts (*IGHA1, IGHG1*) and lactoferrin (*LTF*), while *RORC* mRNA levels, encoding for a T_h_17-specific transcription factor and class II HLA transcripts (*HLA-DMA, HLA-DMB, HLA-DOA, HLA-DPB1, HLA-DPB, HLA-DQA1, HLA-DQB1, HLA-DR1, HLA-DRB1, HLA-DRB5*) were downregulated throughout the disease course ^14,15^. Most of the upregulated transcripts were shared between two or more conditions. Neutrophil marker transcripts (*DEFA3* and *DEFA4)* were particularly elevated in the timepoints with an active inflammatory response (pseudotime 2 to 4).

Next, we employed ImpulseDE2 to construct a model to assess temporal changes in gene expression for individual disease trajectories ^41,42^. Note that this analysis follows a case-only design, exploits intra-individual variation over time and is thus complementary to the pair-wise analysis. Since two of three patients reaching the critical category (pseudotime 2) were deceased soon after reaching this timepoint and did not experience a convalescent disease course, this category was excluded from this analysis and was only used to contrast identified transcripts *post hoc*. We identified 935 differentially expressed genes following impulse-like progression patterns across the disease trajectory (Figure 3d, Supplementary Figure 2a). Genes involved in IL-1β (*IRAK2*, *IL1RN*, *IL1B*, *ICAM1*) and vasodilatory (*AGTRAP*, *ADM*, *NTSR1*, *DM2*, *AOC1*) signalling were highly expressed during the incremental trajectory (pseudotime 1). A broad downregulation of transcripts encoding ribosomal structural proteins was present from the incremental to the late convalescent state (pseudotimes 1,3, and 4), which might reflect a general suppression of the protein synthetic apparatus by type I interferons ^43-45^. Opposingly, there was a strong upregulation of interferon signalling related transcripts (e.g., *OAS1*, *OAS2*, *OAS3*, *IFI44L*, *STAT1*) during the peak of disease activity, which was, however, suppressed in the critical disease category (Figure 3d and Supplementary Figure 2b), confirming earlier reports ^9,10^. Transcripts involved in myeloid cell-mediated immunity and neutrophil degranulation were modestly upregulated during pseudotime 1, 3, and 4, but were strongly increased in the critical pseudotime 2. Notably, during late convalescence (pseudotimes 5 and 6) a strong signal of erythroid cell differentiation (haemoglobin and mitophagy related transcripts, e.g., *HMGN*, *HBD*, *HBBP1*, *OPTN*, *FIS1*) was detectable indicating a response to hypoxia ^46^.

To identify putative transcription factors (TF) responsible for the temporal change of gene expression we used the gene set resource DoRothEA to infer TF activity by enriched regulon analysis. The algorithm accounts for the activity and activity type of the TF downstream targets as well as alternative signalling pathways. Upregulated TFs in active disease were putatively related to inflammation and interferon signalling (*IRF1, STAT3, ESR1*, or *SP1*) as well as hypoxic conditions (HIF1A) (Supplementary Figure 2c). Supporting the IL-21-plasmablast axis identified in the first part of the single-cell analysis, PRDM1 (BLIMP-1) was predicted as significantly active both at the critical as well as at convalescent timepoints. Other TFs with a similar activity pattern are TEAD4, FOXO1, JUNB, SOX2, BACH2, GATA6, RUNX2, THAP11, SOX6, MAFK, and NFE2. A hypergeometric test on these TFs against the REACTOME database showed an enrichment of “cell differentiation pathway” (*q*-value = 0.03) and “factors involved in megakaryocyte development and platelet production” (*q*-value = 0.05). Next, we inferred the upstream network topology of gene expression changes using signed and directed protein–protein interactions ^47^. We reconstructed a network for each pseudotime with a total of 203 unique nodes. To investigate the differences in network topology between pseudotimes, we calculated the degree centralities of every node (Supplementary Figure 2d). MAPK3 (ERK1) and in part MAPK1 (EKR2) has the highest centrality in all pseudotimes (Supplementary Figure 2d). The degree of centrality further indicated how inflammation related nodes, like STAT3 or CTNNB1, a member of the WNT pathway, remain central to the networks across most pseudotimes. Yet, network topology changes between the inflammatory (pseudotimes 1-4) and convalescence (pseudotimes 5-7) states. The former are marked by MAPK14 (p38a) ^48^, ESR1 ^49^ and SMAD4, all regulators of stress and inflammation, while the central nodes of the latter are CREB1, MAPK9, FOXO1, GSK3B, which play a role in reducing inflammation ^50,51^ and vascular remodelling ^52^. In line with this, the number of inferred interactions decreased over time, which suggests substantial normalization of inflammatory signalling networks with recovery. We further constructed metabolic models of each pseudotime using the differentially expressed genes between different disease phases ^53^ and found that the inflammatory disease states are associated with higher numbers of predicted metabolically active pathways when compared to convalescence and healthy individuals (Supplementary Figure 2e). To interrogate the potential presence of viral transcripts, we investigated the bulk RNA data set for presence of viral reads by a stringent mapping approach. We did not detect relevant amounts (>10) of viral reads in any sample. This finding was also confirmed by routine real time-PCR (E gene and S gene amplicon), which did not detect the presence of viral mRNA after 44 cycles in any sample (data not shown).

### Longitudinal co-expression modules identify impaired interferon response in critical disease and signatures of increased thrombo- and erythropoiesis

To identify co-regulated modules of transcription regulated across the disease trajectory of COVID-19, we constructed gene-co-expression networks for all differentially expressed genes identified from the combination of pairwise and longitudinal analysis (6,317 genes) using weighted gene co-expression network analysis (WGCNA) ^54^. Co-expression analysis builds on the hypothesis that genes with similar expression patterns are likely to have a functional relationship or are expressed in specific cell types present in the tissue of interest ^55^. Gene co-expression analysis identified a total of 10 modules, which we refer to as M1-M10, of strongly co-expressed genes following distinct expression patterns throughout the COVID-19 disease phases (Figure 4a and Supplementary Figure 2f). We calculated the eigengene values, which represent a single expression profile for all genes within a module and used these values to assess the individual correlation between the modules and scRNA-seq-derived cellular composition (Figure 4b) and clinical parameters (Figure 4c). Gene set enrichment analysis revealed specific biological processes and pathways that were significantly enriched in each of the modules (Figure 4d) and transcription factor binding site (TFBS) inference ^56^ was used to depict putatively involved transcription factors.

**Figure 4.**
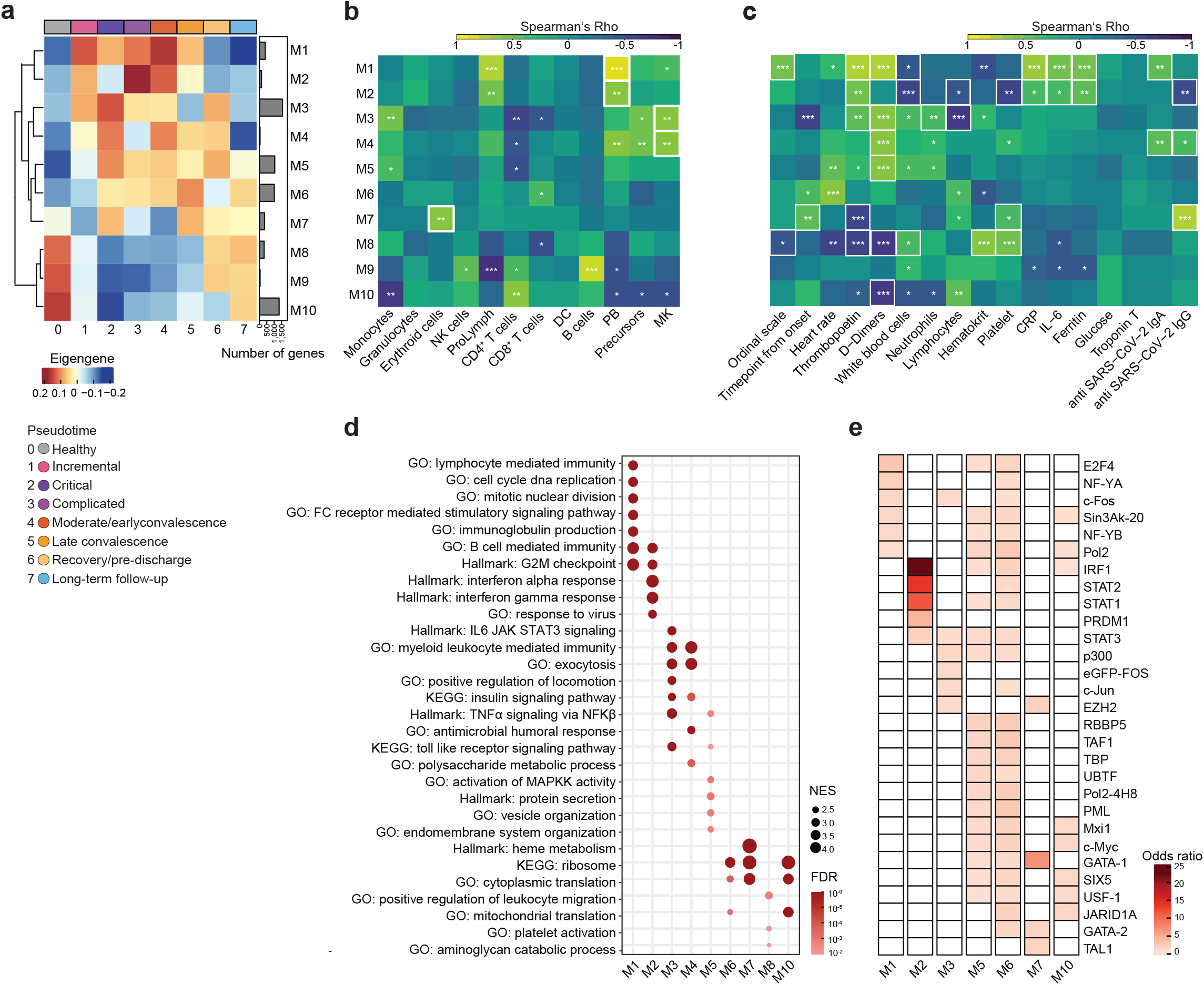
Co-expression analysis of differentially expressed genes. **a**, Group eigengene heatmap of co-expression modules constructed using all pairwise and longitudinal DEGs. The average eigengene values of all samples within each pseudotime are plotted. The number of genes belonging to each module are plotted as a bar plot on the right side of the heatmap. **b**, Correlation heatmap showing Spearman’s rank correlation coefficients between gene co-expression modules (rows) and cell-specific proportion from scRNA-seq data (columns). * indicates *p*-value < 0.05, ** *p*-value < 0.01 and *** *p*-value < 0.001 in Spearman’s correlation. Colour intensity correspond to correlation coefficient. **c**, Correlation heatmap showing Spearman’s rank correlation coefficients between gene co-expression modules (rows) and clinical parameters (columns). * indicates *p*-value < 0.05, ** *p*-value < 0.01 and *** *p*-value < 0.001 in Spearman’s correlation. Colour intensity correspond to correlation coefficient. **d**, Dot plot showing the gene set enrichment analysis (GSEA) of gene co-expression modules against GO (Biological Processes), Hallmark and KEGG gene sets. Size of the dots is proportional to the normalized enrichment score (NES) and the colour corresponds to the false discovery rate (FDR). Selected top terms are visualized. **e**, Heatmap showing the significant enrichment, quantified by odds ratio, of transcription factor binding sites (TFBS) in the gene co-expression modules. Selected top transcription factors are visualized. (See also Supplementary Figure 2)

Genes belonging to modules M1-M7 were upregulated in one or more disease pseudotimes, while those in modules M8-M10 had higher expression levels in the controls and recovery time point (Figure 4a and Supplementary Figure 2f). Module M1, a general inflammation-related module, which was broadly upregulated from pseudotime 1 to 5, is enriched with genes related to lymphocyte mediated immunity, immunoglobulin production and B cell mediated immunity, amongst others (Figures 4a, 4c and 4d). M1 is also highly correlated with the proportion of plasmablasts detected from the scRNA-seq analysis (Figure 4b) as well as D-dimers and CRP levels (Figure 4c). Notable biological processes enriched in module M2 were type I interferon response and proliferative activity (G2M transition) (Figure 4d). Genes belonging to this module were upregulated in all active disease categories except the critical one (Figure 3d). M2 genes are also enriched in binding sites for transcription factors IRF1, STAT1, and STAT2 (Figure 4e). Module M4 was associated with the presence of megakaryocytes in the peripheral blood as well as D-dimers and had two expression peaks, in the critical state (pseudotime 2) and in the early convalescent state (pseudotime 4), respectively. Module M7 contains transcripts related to heme and haemoglobin biosynthesis and coincided with TFBS motifs for GATA-1 and GATA-2, which are classical transcription factors related to erythroid cell differentiation and haematopoietic stem cells. This module, which is likely driven by hypoxic states, had higher levels in the critical phase (pseudotime 2) and was again upregulated in late convalescence (pseudotime 5), which negatively correlates with the prediction of the GATA-2 TF activity (Supplementary Figure 2c). This biphasic pattern is likely related to hypoxia during highly acute inflammation (first peak) and weaning off supplemental oxygen during convalescence (second peak).

Taken together, analysis of temporal gene expression patterns in whole blood of patients clearly depicts pathophysiological consequences of a SARS-CoV-2 virus infection along an idealized disease trajectory.

### Whole blood DNA methylation profiling reveals genome-wide hypomethylation in COVID-19 associated with gene expression

Epigenetic changes have been shown to contribute to the pathophysiology of systemic inflammatory states ^57^. We thus investigated DNA methylation (DNAm) patterns along the COVID-19 disease course following the workflow depicted in Figure 5a. We performed DNA methylation screening on whole-blood samples from a subset of the same patients (*n* = 6) (Table 1) and compared them with a cohort of 6 healthy age and gender-matched controls. Bead arrays interrogating > 850,000 CpG sites across the entire genome covering 99% of RefSeq genes were employed for the analysis. The schematic analysis workflow is depicted in Figure 5a. A pairwise comparison with healthy controls identified between 46,071 and 69,733 differentially methylated CpG sites per pseudotime (Figures 5b and 5d). A general preponderance of hypomethylated sites was present at each timepoint. Intersecting the differentially methylated positions (DMPs) between pseudotimes, we noted that most sites were unique to a respective category. Among the top 30,000 DMPs, 3804 DMPs were shared among the first three timepoints (pseudotimes 1, 2, and 3), which are clinically characterized by strong systemic inflammation. 875 DMPs were common between the last three pseudotimes, which reflect the convalescence trajectory with decreasing disease activity (Figure 5c). Of note, the analysis represents the sum signals of the DNA methylome in peripheral blood and thus many of the DMPs may actually reflect cell type-specific methylation patterns. A formal cellular deconvolution analysis using RnBeads ^58,59^ identified that major parts of the COVID-19 associated DNAm signatures originated from granulocytes, B cells, NK cells and monocytes (Supplementary Figure 3a). Next, we analyzed the enrichment of TF binding motifs within the regions containing differentially methylated CpG sites across timepoints using Locus Overlap Analysis (LOLA)^60^. In the inflammatory disease phase and again late in disease, we observed a significant overrepresentation of binding sites of the CCAAT Enhancer Binding Protein Beta (CEBPB) in hypomethylated regions (Figure 5e), which is a transcription factor present in activated monocytes ^61^ and in T and NK cells. CEPBP has a critical role for emergency granulopoiesis ^62^. Interestingly, high levels of CEBPB activity have also been associated with B lymphocyte-to-granulocyte trans-differentiation, a process, which has been proposed in severe COVID-19 ^15^. Binding motifs for BATF, a cooperative transcription with heterologous interaction partners (e.g., BACH2 and IRFs) were strongly enriched at hypermethylated sites in the critical phase (pseudotime 2) and late during convalescence, indicating a potential antagonization of the function of this TF. The BATF family has several pleiotropic functions in adaptive immunity, e.g., regulating T_h_17 or T_h_2 immunity as well as promoting class switching in B cells ^63-65^.

**Figure 5.**
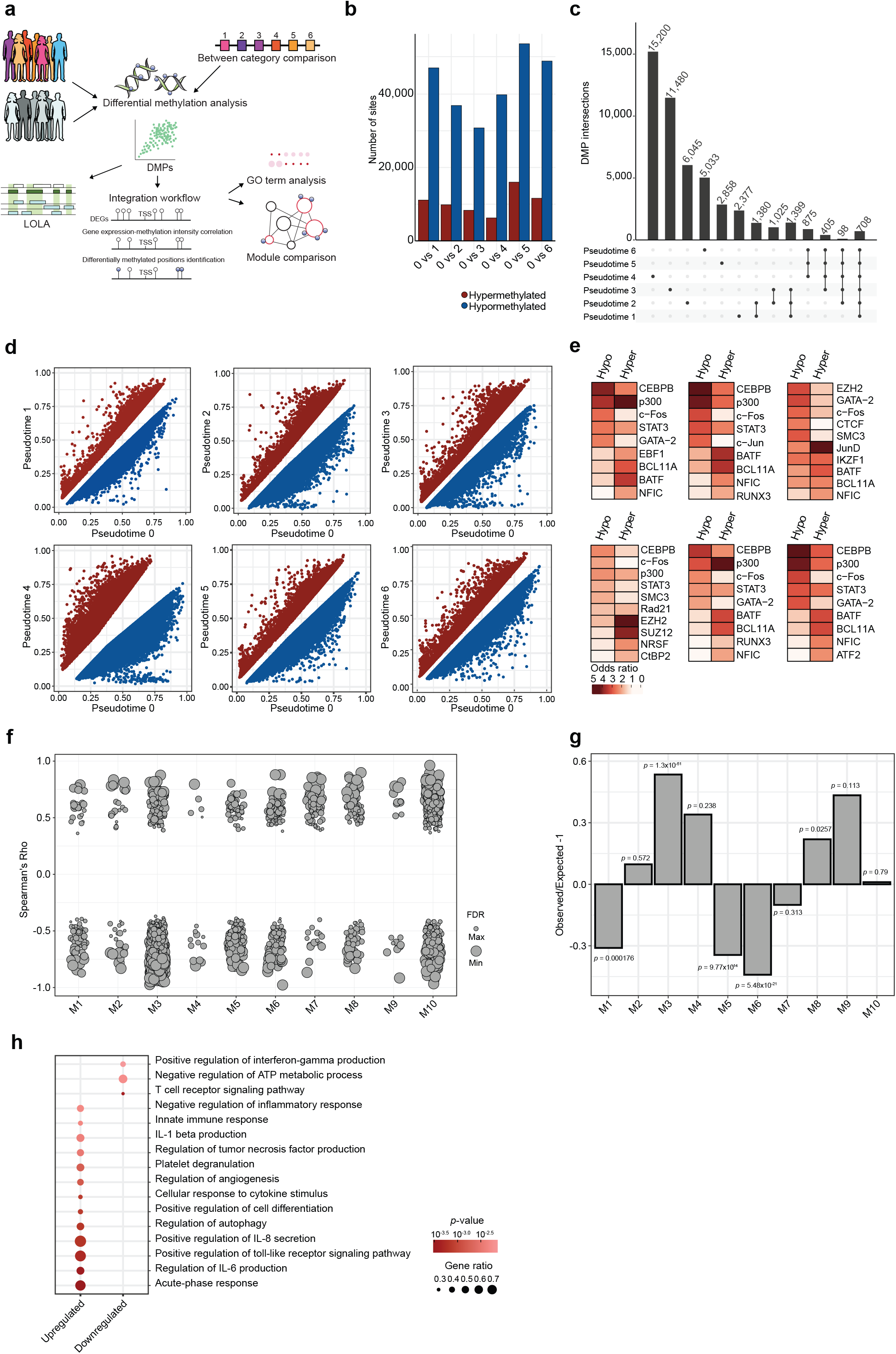
Genome-wide DNA methylation patterns in COVID-19. **a**, Schematic workflow of the analysis performed on the whole blood EPIC array data and its integration with bulk RNA-seq data. **b**, Number of significantly differentially methylated positions (DMPs) between controls and each of the COVID-19 pseudotimes. Colours discriminate hypermethylated (red) and hypomethylated (blue) positions in COVID-19 samples compared to controls. **c**, DMP comparisons between top 30,000 DMPs at each pseudotime. Vertical bar plots indicate the number of DMPs specific to each pseudotime (left) by several pseudotimes (right), whereby the contributing groups are indicated as connected dotes (bottom). Only selected overlaps are visualized. **d**, Comparison of mean methylation intensities (β-values) between COVID-19 pseudotimes and healthy controls. Significant DMPs, defined by the automatically selected rank cutoff by RnBeads, are highlighted in red (hypermethylated) and blue (hypomethylated). **e**, Heatmap showing the significant enrichment, quantified by odds ratio, of transcription factor binding sites (TFBS) in the DMPs identified at different COVID-19 pseudotimes. Order of the heatmaps correspond to the comparison depicted in panel **d**. Selected top transcription factors are visualized. **f**, Correlation between co-expression module genes and their nearby DMPs. Each point represents a gene and the size of the points is proportional to statistical significance (FDR) of the correlation with larger points being more significant. DMP-DEG correlations with FDR < 0.05 are visualized. The positions of individual points are jittered horizontally in order to show the density of the data. **g**, Over-representation and under-representation of significantly correlated DMP-DEG in each of the co-expression modules. The over-/under-representation is quantified as the ratio of observed and expected number of correlated genes present in each module under the Chi-square distribution. **h**, Dot plot showing GO terms enriched in upregulated and downregulated DMP-DEG pairs. Size of the dots is proportional to the gene ratio and the colour corresponds to the *p*-value of the enrichment. Selected top terms are visualized. (See also Supplementary Figure 3)

In addition to the pairwise analysis, we also compared methylation patterns between subsequent pseudotimes in order to identify the changes in methylation occurring along the disease trajectory. While we did not observe significantly differentially methylated sites between the timepoints with strong systemic inflammation (pseudotime 1, 2, 3), up to 50,876 sites were differentially methylated during convalescence (Supplementary Figure 3b and 3c). In contrast to the early timepoints, the majority of the DMPs were hypermethylated during convalescence (Supplementary Figure 3b and 3c). TFBS enrichment analysis revealed that binding sites for CEBPB were enriched in hypermethylated sites in pseudotime 6 while that for BATF were enriched in hypomethylated sites in pseudotime 6, suggesting a gradual “restitution” of the healthy methylation state (Supplementary Figure 3d).

Subsequently, we integrated the DEG dataset with the observed DNA methylome changes to identify mRNA expression differences associated with *cis*-linked methylation changes. We hypothesized that this approach would reveal another layer of important genes as DMP-DEG pairs may represent a potentially longer-term modulation of cellular programs or differentiation events. We employed a hierarchical testing approach ^66^ to identify interactions between the COVID19-dependent alterations in the transcriptome and DNA methylation signatures (Figure 5a). To that end, we screened all differentially expressed genes (combined set) for DMPs within a 5kb window up- and downstream of their respective transcription start site. Of all 3,280 DMP-DEG pairs 2,240 (68.3%) showed increased expression with reduced methylation or decreased expression with increased methylation, whereas 1,040 pairs (31.7%) did not follow the canonical inverse pattern, but were positively correlated, which is in line with previous studies ^67^ (Supplementary Figure 3e). We next investigated the representation of DMP-DEG pairs in the longitudinal co-expression modules M1 to M10. Rank-based correlation analysis identified the presence of *cis*-linked DMP-DEG pairs in all modules, with a significant overrepresentation in M3 and M8, whereas DNAm-linked expression changes were significantly depleted from M1, M5, and M6 (Figure 5f and 5g). DMP-DEG pair genes which were either upregulated in the inflammatory phase (pseudotime 1-4) or downregulated in late convalescence (pseudotime 5 and 6) were mapped to scRNA-seq data, identifying larger cell type specific clusters of DMP-DEG pairs for plasmablasts, monocytes, megakaryocytes (Supplementary Figures 3f and 3g).

GO term enrichment analysis of DMP-DEG pairs showed a significant enrichment of innate-immunity related terms in the upregulated DEGs (e.g. TNF and IL-6 signalling, TLR pathway), and also identified gene sets related to platelet function and metabolic processes (ATP metabolism and autophagy) to be significantly altered (Figure 5h). Notable genes from this analysis that were differentially methylated and expressed along the COVID-19 disease trajectory were genes involved in metabolism and autophagy, such as *HK2, PPIF, DDIT4, XBP1*, *ULK1*, *OPTN*, and *PINK1*.

In line with previous findings in other systemic inflammatory states such as sepsis, our analysis pinpoints several immune-related processes to be potentially controlled by DNA methylation changes upon systemic inflammatory responses ^57^ and highlights the link of cellular immunometabolism and epigenetic alterations ^68^.

As megakaryocytes and plasmablasts presented with unexpectedly large clusters of DMP-DEGs, we decided to focus on these cell types in greater depth.

### Plasmablast and B cell changes across the COVID-19 disease trajectory

Humoral immune responses are known to have a major role in virus-induced immunopathologies and have been shown to pivotally contribute to systemic inflammatory responses^69^. Secreted antibodies may have beneficial (e.g. virus neutralization), but also detrimental functions, e.g., hyperactive complement fixation and activation. Several reports of severe systemic inflammatory states, such as Dengue fever and malaria, have highlighted the massive expansion of antibody-producing plasmablasts in the hyperinflammatory phase ^70^. These cells are known to secrete immunomodulatory cytokines like IL-10, may form an immunometabolic sink for nutrients and might thus be an important part of a virus-induced immune dysregulation. Initial studies have shown an expansion of plasmablasts in COVID-19 ^15,71^. However, little is known about the dynamics and functional states of these cells during SARS-CoV-2 infection.

As our findings pointed to a dynamic regulation of the B cell compartment along the disease trajectory of COVID-19, we thus investigated the changes of B cells and plasmablasts in greater detail (Figure 6a). We first extracted 22,190 cells identified as part of the B cell lineage from scRNA-seq data in cohort 1. UMAP embedding of this subset of cells identified two distinct large clusters reflecting *bona fide* B cells, which included 11,509 memory B cells, 3,993 naïve B cells, 383 transitional B cells, and 6,295 plasmablasts (Figure 6b).

**Figure 6.**
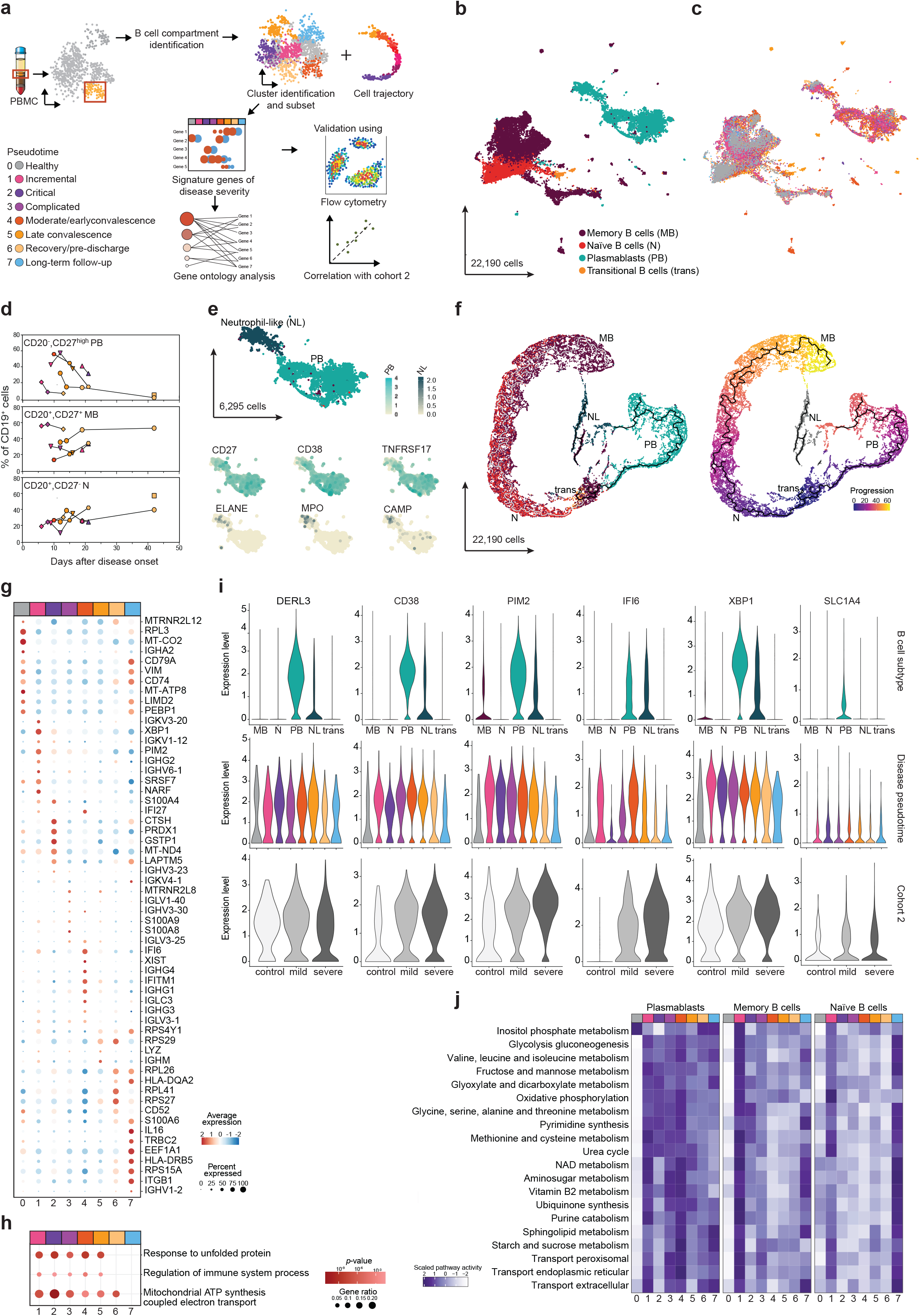
B cell compartment scRNA-seq analysis and flow cytometry identifies plasmablast changes across the COVID-19 disease trajectory. **a**, Schematic workflow of the scRNA-seq analyses performed on the B cell compartment identified in Figure 2. **b**, B cell compartment subtypes represented as a UMAP. In total, 22,190 cells are depicted. Clusters represented are Memory B cells (MB, dark red), naïve B cells (N, red), transitional B cells (trans-orange) and plasmablast (PB, blue). **c**, B cell compartment pseudotimes represented as a UMAP. **d**, Flow cytometry analysis of B cell subtypes. CD19^+^ B cells were stained for CD20 and CD27. CD20^-^CD27^high^ B cells classified as PB, CD20^+^CD27^+^ cells as MB and CD20^+^CD27^-^cells as N. Proportions of each cell type among CD19^+^ B cells were plotted against the sampling time relative to the disease onset. The points were coloured by corresponding pseudotime and connected by patient. Only patients from Kiel cohort (*n* = 7 individuals) are shown. **e**, Plasmablast-specific UMAP highlighted neutrophil-like cells (NL). Smaller UMAPs corresponding to expression of plasmablast markers (*CD27*, *CD38*, and *TNFRSF17*) and neutrophil-like markers (*ELANE*, *MPO*, and *CAMP*). **f**, Cell trajectory analysis of B cell compartment. Cell trajectory was calculated using Monocle3 and B cell subtypes are displayed in left panel followed by the corresponding transition-pseudotime analysis rooted on transitional B cells (purple) and differentiated into 2 branches: B cells naïve/memory branch (grey line/culminating in yellow) and an over imposed plasmablast branch, from which most of the plasmablasts (black line/culminating in orange) are not connected to neutrophil-like cells (grey). **g**, Dot plot for pseudotime signature genes in plasmablasts. Genes were selected based on ten most characteristic upregulated genes. Colour discriminates upregulated (red) or downregulated (blue) genes, while point sizes represents the number of cells per group expressing the corresponding gene. **h**, GO enrichment analysis for upregulated genes during disease trajectory. Size of the dots is proportional to the gene ratio and the colour corresponds to the *p*-value of the enrichment. Selected top terms are visualized. **i**, Gene expression of genes of interest in B cell subtypes, pseudotimes and cohort 2. Genes of interest selected based on their high expression in PB or NL and upregulated during disease (*DERL3*, CD38, *PIM2, IFI6*, *XBP1*, and *SLC1A4*). For each gene, top violin plot depicting B cell subtypes expression, centre violin plot based on pseudotime expressions and bottom violin plot based on the expression of cohort 2 divided by disease classification, healthy control (white), mild disease (light grey) and severe disease (dark grey). **j**, Metabolic pathways enriched in B cell compartment subtypes. Top 20 active metabolic pathways for context-specific metabolic networks reconstructed in plasmablasts are shown. For each B cell subtype, significant differences in metabolic activity were determined using a Kruskal-Wallis test. Number of reaction counts found per pathways is displayed as colour intensity.

Similar to a previous study (Wilk et al., 2020), plasmablasts were largely expanded during COVID-19 with highest levels in the hyperinflammatory phase (Figure 6c). Multicolour flow cytometry analysis confirmed extremely high levels of circulating CD27^hi^CD20^-^ cells in the fraction of CD19^+^ cells, which dropped along the disease convalescence. Likewise, the early relative decrease of naïve (CD20^+^CD27^-^) and memory B cell (CD20^+^CD27^+^) levels normalized at later timepoints (Figure 6d). We also identified a large fraction of HLA-DR^+^CD138^+^ double positive plasmablasts in the inflammatory pseudotimes (pseudotimes 1 to 4) that are usually not present in the peripheral blood (Supplementary Figure 4c). Whereas CD138^+^ cells were only abundant in the active disease and not in the convalescence phase, 82-98% of all CD27^hi^CD20^-^ cells remained HLA-DR positive (Supplementary Figure 4c). This finding is remarkable, as plasmablasts usually lose their ability for antigen presentation during maturation and gain CD138 expression. In healthy controls, 44.72% (SD= 7.87) of all B cells were CD138^+^ plasmablasts and 64.74% (SD= 12.06) HLA-DR^+^ plasma cells (data not shown). Of note, as plasmablasts are notoriously sensitive to manipulation, we recognized that the freezing procedure for scRNA-seq slightly diminished the levels of intact plasmablasts compared to FACS data. Also, corresponding to flow cytometry data, we found similar expression patterns of marker genes for plasmablasts and/or plasma cells (*CD27*, *CD38*, *MZB1*, *SLAMF7*, *SDC1* and *HLA-DR*), which hindered the distinction of the two cell types, prompting us to use the overarching term plasmablasts. Regardless, the longitudinal dynamics of increased plasmablast numbers were retained between the two methods.

Projected in direct vicinity to conventional plasmablasts, we could distinguish a smaller cluster which expressed genes normally associated with neutrophils (e.g., *ELANE*, *MPO* and *CAMP*) (Figure 6e) ^15^. Unlike in the previous study, we also found such cells in healthy subjects, although at lower frequency (Supplementary Figure 4b). Constructing a cell differentiation trajectory based on transcriptional reconfiguration of B cells using monocle3 ^72^, we highlighted the progression of transitional B cells into naïve and memory B cells, with a separate memory B cell cluster being most likely CD45RB^-^cells ^73^ (Figure 6f). In parallel, we could retrace the progression of plasmablasts, which did not cluster based on immunoglobulin classes but rather based on pseudotimes with cells from hyperinflammatory phases present further away from the root of the cell trajectory (Supplementary Figures 4a and 4b). Interestingly, cells previously identified as neutrophil-like cells associated to the plasmablast compartment were not continuous within the plasmablast differentiation (Figure 6f).

We next turned to the analysis of BCR repertoire in the different B cell compartments using heavy-chain bulk- and scBCR-seq (Supplementary Figures 4d-o). In the bulk data set, we identified all together 596,882 unique BCR CDR3 heavy-chain sequences, while for scBCR-seq we had information referring to 14,785 cells. Diversity analysis showed a heightened clonality, which sharply increased at early timepoints and then gradually decreased in the convalescent pseudotimes until normalization in follow-up (Supplementary Figure 4d). A similar pattern was also discerned in plasmablasts although not so conspicuous (Supplementary Figure 4e). Bulk BCR identified an expansion of IgA^+^ and IgG^+^ cells (Supplementary Figures 4f and 4h), with the most frequently used IGH genes -*IGHA1* and *IGHG1*-expression increased during COVID-19 in both memory B cells and plasmablasts (Supplementary Figures 4g and 4i). Of note, *IGHA1* and *IGHG1* increased expression was reached earlier in plasmablasts compared to memory B cells, while *IGHA2* and *IGHG2* expression was only distinguishable in plasmablasts. Despite lower numbers, for scBCR-seq we could still distinguish expanded IgA^+^ and IgG^+^ cells upon disease recovery, mostly driven by plasmablasts (Supplementary Figures 4j and 4m). To study the preponderance of certain V gene regions in COVID-19 patients, we analysed the usage of IGHV family subunits in combination with specific IGHA and IGHG genes (Supplementary Figures 4k and 4n). For *IGHA1*, the V gene regions *IGHV3-23*, *IGHV1-18*, *IGHV4-59*, and *IGHV4-39* were preferentially associated in COVID-19 patients compared to controls. For *IGHG1* expressing cells, the preferred IGHVs were *IGHV3-30*, *IGHV4-39*, *IGHV1-2*, and *IGHV3-23*. Both *IGHV3-30* and *IGHV3-23* were present in plasmablasts and neutrophil-like cells during disease (Supplementary Figures 4l and 4o). In summary, we observed an increase in B cell clonality in COVID-19, with a massive increase of memory B cells and plasmablasts, which was dominated by the IgA and IgG isotypes - typical for mucosal immune reactions - and a skewed use of the *IGHV* gene early during the disease course.

As we hypothesized that plasmablasts are important immunological players during SARS-CoV-2 infection, we next analysed the longitudinal gene expression patterns of the 6,295 plasmablasts along the disease trajectory. Indeed, significant changes pointed to a shift of pathways and cellular programs across timepoints (Figure 6g). Whereas in circulating plasmablasts from healthy controls, transcripts related to early immunoglobulin production (*IGHA2*), BCR signalling (*CD79A*) and MHC-II maturation prevailed, the incremental inflammatory phase of COVID-19 patients was characterized by ER stress/protein folding (*XBP1*) and cell proliferation (*PIM2*, *S100A4* and *NARF*). Type I IFN response genes (*IFI27*, *IFI6*, *IFITM1*) were present in the plasmablasts along the trajectory until late in the disease (up to pseudotime 5), yet such genes were absent from plasmablasts in critically ill patients (pseudotime 2). The convalescence and recovery states (pseudotimes 5, 6 and 7) were characterized by upregulation of HLA class II (*HLA-DQA2* and *HLA-DRB5*) and upregulation of *CD52*, which is known to be expressed on specific activated B cells (ABCs), that are committed to the memory B cell lineage ^74^. Throughout disease we also identified an upregulation of *SLC1A4*, a neutral amino acid transporter and a potential upstream regulator of metabolic changes (Figure 6i). IL-16 secretion, which has been shown to support migration of CD4^+^ T cells ^75^ and circulating blood DCs into lymphoid organs during the initiation of a humoral immune response is a feature of late plasmablasts (Figure 6g). GO enrichment analysis identified unfolded protein response and mitochondrial ATP synthesis as upregulated processes during active disease (Figure 6h). These findings were also corroborated for plasmablasts from an independent cohort (cohort 2) of mild and severe COVID-19 patients using another scRNA-seq technology (Rhapsody^TM^ BD) (Schulte-Schrepping et al., 2020). 2,263 plasmablasts extracted from this dataset revealed that those of severe patients also displayed an increase expression of *CD38*, *PIM2*, *IFI6*, *XBP1*, and *SLC1A4* and similarly enriched GO terms (Fisher test, *p* = 1.80×10^-5^ for unfolded protein response and *p* = 2.50×10^-8^ for mitochondrial ATP synthesis).

As metabolically active cells, plasmablasts have been reported to modulate immune responses by serving as a nutrient sink ^76^. Thus, we next applied our cell-specific metabolic model to the expression data of cell-types of interest, using a constraint-based method through the reconstruction of the metabolic state of individual cells from scRNA-seq data core reactions ^77^. Plasmablasts found in inflammatory states of the disease trajectory (pseudotimes 1-4) were associated with metabolic hyperactivity, which was reduced only upon recovery to the state of healthy subjects (Supplementary Figure 6a). While memory and naïve B cells displayed no significant differences in overall metabolic activity between disease states (Supplementary Figure 6a). Predicted metabolic processes related to plasmablasts activity increase were identified as an induction of oxidative phosphorylation, glyoxylate and dicarboxylate metabolism, NAD synthesis, an increase of various amino acid metabolic pathways (including glycine, serine, alanine, histidine, threonine, glutamine and arginine) and an upregulation of several amino acid transporter pathways. Glycolysis is predicted to have a low activity state in inflammatory disease phases, whereas it is highly active at clinical recovery (pseudotime 6) (Figure 6j).

Taken together, the analysis identifies the broad activation of plasmablasts and suggests a strong immunometabolic shift of the cells towards amino acid catabolism, which may contribute to the immunopathology seen in severe COVID-19.

### Elevated megakaryocyte levels as a feature of the systemic inflammatory response to COVID-19

Systemic inflammatory responses are known to consume platelets, which exert broad immune and inflammatory functions in addition to their well-established haemostatic role ^78,79^. The interaction of platelets with neutrophils orchestrates the formation of neutrophil extracellular traps ^79,80^, which elicits a rapid depletion of platelets from the circulation, resulting in a transient thrombocytopenia ^81,82^. Interestingly, platelet-rich thrombi and increased megakaryocytes were found in the lungs of deceased patients ^28^ and pulmonary and cerebral embolism is an important contributor to morbidity and mortality in COVID-19 ^83^. As we had observed a transient increase of circulating megakaryocytes, the cellular source of platelets (Figure 2b), in the single-cell data and identified co-expression modules (M3 and M4) related to platelet degranulation and D-dimer levels, we hypothesized that altered presence and function of megakaryocytes might be a distinct feature of COVID-19. We thus performed sub-clustering of 6,512 cells identified as megakaryocytes and their respective haematopoietic stem cell precursors (HSCs and Megakaryocyte-Erythroid precursors, MEPs) using *k*-nearest neighbour method (Figure 7a and Supplementary Figure 5a). The cells clustered into two distinct subgroups, 5,870 identified as *bona fide* megakaryocytes and another smaller cluster containing all HSCs and MEPs (Figure 7b and Supplementary Figure 5b). The relatively low number of cell precursors complicated the comparison between individual pseudotimes (Supplementary Figures 5d-g). However, we could discern a significant increase of HSCs and MEPs at the convalescence state in comparison with healthy controls (Supplementary Figure 5j).

**Figure 7.**
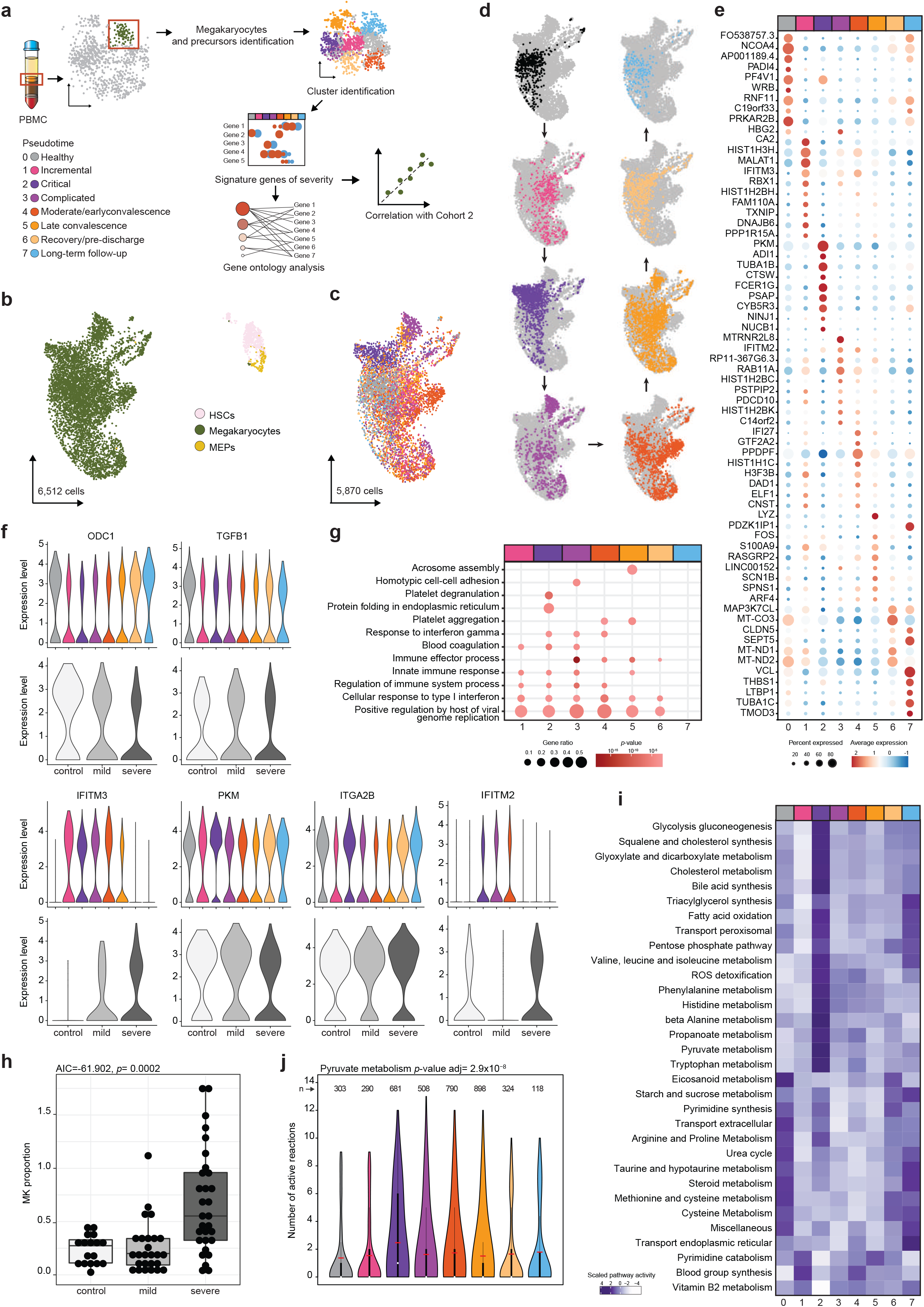
Elevated megakaryocyte levels as a feature of the systemic inflammatory response to COVID-19. **a**, Schematic workflow of the scRNA-seq analyses performed on the megakaryocytes identified in Figure 2. **b**, Megakaryocytes and their precursors as a UMAP. In total 6,512 cells are depicted. Clusters represented are Megakaryocytes (green), HSCs (pink) and MEPs (yellow). **c**, Megakaryocytes pseudotimes represented as a UMAP. In total, 5,870 cells are depicted. **d**, Megakaryocytes across COVID-19 disease trajectory. For each UMAP, pseudotime-specific cells were highlighted by colour. **e**, Dot plot for disease trajectory signature genes in megakaryocytes. Genes were selected based on ten most characteristic upregulated genes. Colour discriminates upregulated (red) or downregulated (blue) genes, while point sizes represents the number of cells per group expressing the corresponding gene. **f**, Gene expression of genes of interest across pseudotimes and cohort 2. Genes of interest included genes that were downregulated during disease (*ODC1* and *TGFB1*) and genes the are upregulated during disease (*IFITM3*, *PKM*, *ITGA2B*, and *IFITM2*). For each gene, top violin plot based on pseudotime expressions and bottom violin plot based on the expression of cohort 2 divided by disease classification, healthy control (white), mild disease (light grey) and severe disease (dark grey). **g**, GO enrichment analysis for upregulated genes during disease trajectory. Size of the dots is proportional to the gene ratio and the colour corresponds to the *p*-value of the enrichment. Selected top terms are visualized. **h**, Cohort 2 megakaryocyte proportions grouped by disease severity. Healthy control (white), mild disease (light grey) and severe disease (dark grey) were depicted. AIC and *p*-value based on linear mixed model. **i**, Metabolic pathways enriched in megakaryocytes. Top differentially active metabolic pathways for context-specific metabolic networks reconstructed are shown. Significant differences in metabolic activity were determined using a Kruskal-Wallis test. Number of reaction counts found per pathways are displayed as colour intensity. **j**, Pyruvate metabolism in megakaryocytes. Number of pyruvate metabolic pathway active reactions by pseudotimes were depicted. *p*-value based on Kruskal-Wallis test. Number of models per pseudotime were denoted as n above each column.

By focusing exclusively on megakaryocytes there was a clear separation between samples belonging to healthy controls and patients with active COVID-19 (Figure 7c), particularly cells from complicated disease phases form distinct subclusters from cells of healthy and long-term recovery disease phases (Figure 7d). Differential gene expression analysis between healthy controls and the different disease pseudotimes identified patterns of transcripts being upregulated along the disease trajectory and diffusion maps showed a distinct subclustering of megakaryocytes (Figures 7d and 7e). Notably top 10 indicator transcripts of the incremental inflammatory phase (pseudotime 1) include *IFITM3*, encoding an interferon-induced membrane protein that helps confer immunity against viruses and *MALAT1*, a lncRNA, which serves as a transcriptional regulator for genes involved in cell migration and cell cycle regulation. Strong upregulation of PKM (*PKM2*), a pyruvate kinase that is involved in ATP formation, interacts with HIF1A and promotes its activity, was observed in critical patients (pseudotime 2). In this group, high expression of *FCER1G*, encoding the common FcR γ-chain adaptor responsible for integrin (ITGA2/GP6)-mediated platelet adhesion was also present (Figure 7e). Other interferon response genes were among the top 10 differentially expressed genes, such as *IFITM2* and *IFI27* genes involved in interferon gamma pathway. gene ontology (GO) enrichment analysis. Thus, upregulation of broad terms related to immune responses, type I IFN response and platelet aggregation along the disease trajectory were identified in GO enrichment analysis (Figure 7g).

Transiently downregulated transcripts comprised of *ODC1* (ornithine decarboxylase), the rate-limiting enzyme for polyamine synthesis ^84^, which is known to inhibit platelet aggregation ^85^, and *TGFB1* (Figure 7f). Of note, low levels of *TGFB1* in megakaryocytes are involved promoting a regenerative state of HSCs in the bone marrow ^86^. *TREML1* and *ITGA2B* (*CD41*) expression as canonical marker transcripts for megakaryocytes and platelets ^87^ remained rather stable throughout the disease course with a slight upregulation in the critical phase. *IFITM3*, *IFI27*, and *IFITM2*, which were shown to be induced in emergency megakaryopoiesis, were upregulated in the inflammatory pseudotimes 1, 2, 3, and 4, indicating a lasting interferon response throughout the disease trajectory in megakaryocytes. Furthermore, we validated our findings in an independent cohort of mild and severe COVID-19 ^14^, which confirmed not only higher numbers of megakaryocytes in severe patients (Figure 7h), but also confirmed the upregulation of *IFITM3*, *PKM*, *ITGA2B* (CD41), and *IFITM2*, and the downregulation of *ODC1* and *TGFB1* in the severely ill patients (Figure 7f).

We next employed our metabolic modelling approach to depict active metabolic pathways in megakaryocytes. They showed increased metabolic activity along the disease trajectory compared to healthy patients, albeit at a lower level compared to plasmablasts (Figure 7i). Notable predicted processes were related to energy metabolism (pyruvate metabolism, glycolysis, and ROS detoxification). Metabolic plasticity related to these processes, but also a direct function of pyruvate kinase M in PI3K/Akt signalling has been shown to enhance platelet aggregation ^88,89^ (Figure 7j and Supplementary Figure 6c). Focusing on metabolic changes in central metabolism based on metabolic modelling, we observed a pronounced up-regulation of glycolytic flux toward lactate which has been associated with increased platelet aggregation ^89^. This was also evidenced by a pronounced induction of methylglyoxal degradation pathways with methylglyoxal being a side-product of a highly active glycolysis ^90^ and itself causing platelet hyperaggregation (Hadas et al., 2013). Together with the downregulation of spermidine products known to inhibit platelet aggregation and induction of spermidine degradation ^85^ (Supplementary Figure 6b).

Taken together, our results demonstrate a significant upregulation of megakaryocytes in the peripheral circulation of COVID-19 patients. These megakaryocytes show distinct temporal gene expression patterns along the disease trajectory with clear features of interferon-induced emergency megakaryopoiesis, which has been linked to functional exhaustion of megakaryocyte progenitors. This functional state of megakaryocytes may be involved in the hyper-aggregative state of platelets in the COVID-19, thrombocytopenia, and may ultimately foster micro- and macrothrombotic complications, which are a hallmark of severe COVID-19 disease courses.

### Interrogating co-expression modules in a larger longitudinal cohort of severe COVID-19 patients confirm clinical significance of skewed cellular programs

Lastly, we focused on the potential importance of the obtained signatures in a clinical context in a longitudinal cohort of 40 mechanically ventilated, critically ill COVID-19 patients from Radboud University Medical Center (UMC) in Nijmegen (cohort 3) (Supplementary Table 3). We first sought to correlate the levels of previously defined co-expression modules (see Figure 4a) linked to distinct dysregulation features of peripheral blood cells with clinical outcome. Bulk RNA sequencing data in this cohort were obtained at two timepoints early upon admission to the ICU. We used matched sample pairs with similar increasing inflammatory activity changes from survivors (*n* = 33) and non-survivors (*n* = 7) and interrogated the change of expression levels of module genes (M1 to M10, as defined in the German longitudinal cohort (cohort 1) initially shown in Figure 4a) between the two timepoints (Figure 8a). The first sample was obtained in median 3 days after ICU admission, the median period between the time points was 2 days without systematic differences between surviving and non-surviving patients. The second timepoint of non-survivors varied between 4-35 days before death. Three modules were significantly regulated in this longitudinal comparison. M2 transcripts related to failing type I interferon response and proliferative activity (G2M transition) were significantly downregulated in both survivors and non-survivors, corroborating the association of interferon dysregulation and severe disease ^9,10,69^. M4, indicative of megakaryocytes in the peripheral blood as well as elevated D-dimer levels and M7 transcripts, associated with erythroid differentiation and megakaryocytes, were significantly upregulated only in COVID-19 non-survivors when comparing the change between sampling at ICU admission and the follow-up time point two days later (Figure 8b).

**Figure 8.**
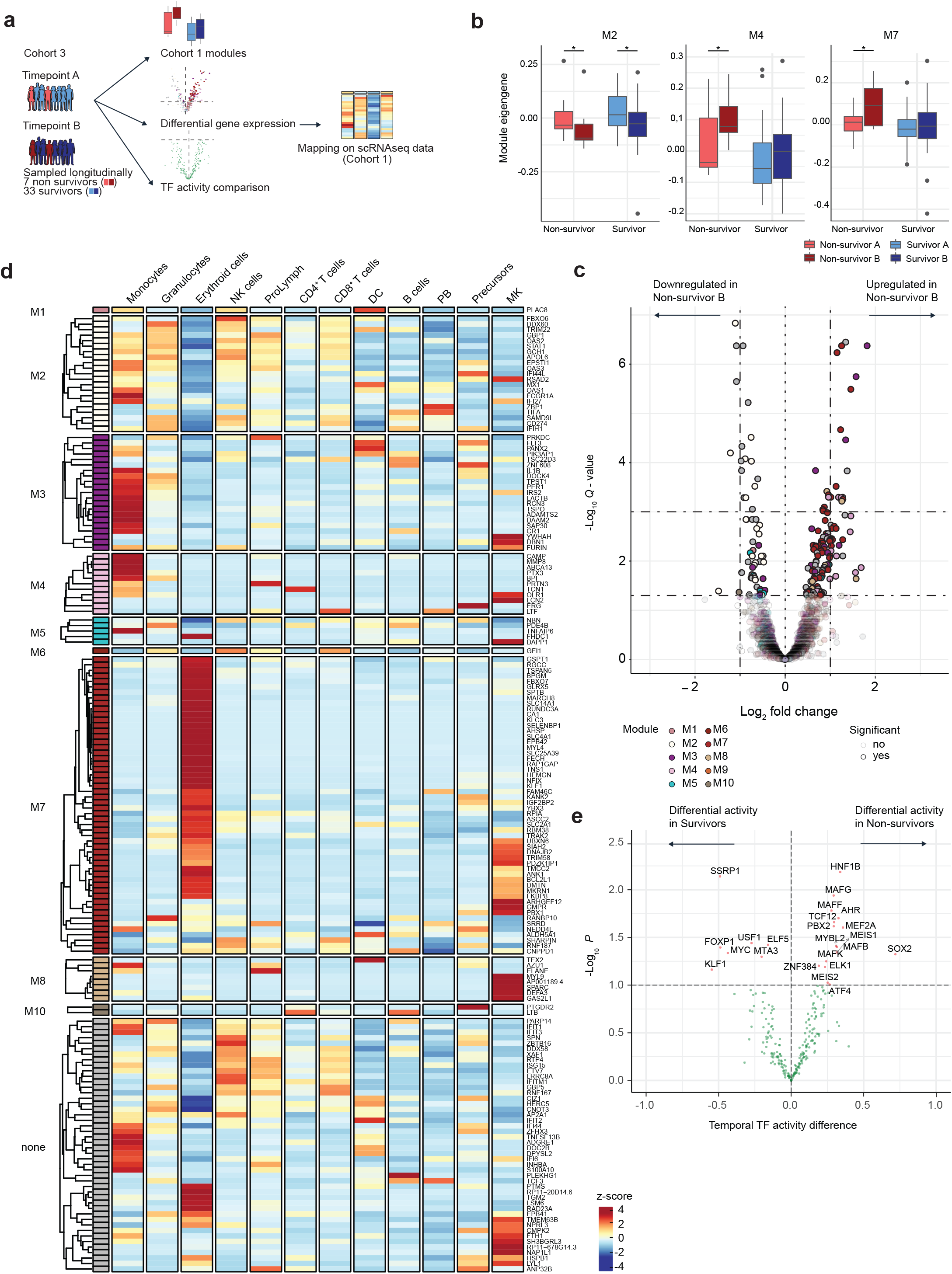
Interrogating co-expression modules in a larger longitudinal cohort of severe COVID-19 patients confirm clinical significance of skewed cellular programs. **a**, Schematic workflow of the analysis performed on bulk RNA-seq data from cohort 3. **b**, Module eigengene comparisons between the two sampled timepoints in survivors and non-survivors for M2, M4 and M7. **c**, Volcano plot depicting log2-fold changes and FDR adjusted *p*-values between the two sampled timepoints in non-survivors. Genes are colour-coded by the corresponding co-expression modules. Darker colours represent significantly differentially expressed genes. **d**, Heatmap showing the average expression of differentially expressed genes identified in non-survivors in different cell-types of cohort 1 (from scRNA-seq data). The average expression in severe stages of the disease (pseudotimes 1, 2, and 3) is shown. The average genes counts are scaled by row and row-wise *z*-scores are plotted in the heatmap. Genes are labelled by the co-expression modules and are hierarchically clustered for each module separately. **e**, Volcano plot depicting the differential Transcription factor activity over time in non-survivors (right) vs. survivors (left) versus the -log10 transformed p-value. Significant TFs (p-value < 0.1) are marked in red and annotated with the gene symbol.

In a reverse approach, we next asked which transcripts displayed longitudinally different expression patterns between the two timepoints in survivors vs. non-survivors. We found that while in survivors only 3 transcripts were regulated, 182 transcripts were significantly different between the first and the second time point (Figure 8c). Of these transcripts, 130 were contained in the previously defined temporal co-expression modules from cohort 1 with a significant enrichment of transcripts related to M2, M4, and M7. To investigate cell-type specific expression patterns of these fatality-associated transcripts, we plotted their respective average expression values for each identified cell population from the inflammatory states from cohort 1. We observed distinct clusters of genes which had high expression levels in specific cell types. While the expression signature for M2 genes marked a broad array of cell types, such as monocytes, granulocytes, NK cells, proliferative lymphocytes and CD8^+^ T cells, M3 and M4 genes were mostly specific for monocytes, with few highly expressed transcripts attributed to megakaryocytes. A large proportion of M7 genes painted the erythroid lineage, a separate cluster was expressed specifically in megakaryocytes, e.g., *PBX1*, *GMPR*, *ARHGEF12*, *TRIM58*, and *PDZK1IP1* (Figure 8d).

Similar to the transcriptional patterns, predicted TF activities of the two cohorts were highly correlated for the inflammatory disease states (Permutation test, *p*-value < 10^-16^). Lastly, we quantified the differential regulation of transcription factors over time in survivors vs. non-survivors via a moderated t-test using limma ^91^, while accounting for patient correlation. The analysis identified 16 TFs that were differentially regulated in non-survivors only and 7 TFs in survivors (Figure 8e). Pathway analysis (REACTOME) revealed significant enrichment of the terms “factors involved in megakaryocyte development and platelet production” (p-value = 0.0001) and “TRAF6 mediated induction of proinflammatory cytokines” (p-value = 0.001) in the non-survivor TFs, while pathways such as “TFAP2 (AP-2) family regulates transcription of cell cycle factors” (p=0.002), “binding of TCF/LEF:CTNNB1 to target gene promoters” (p=0.003) and “RUNX3 regulates WNT signaling” (p=0.003) were enriched in survivor TFs. Collectively, the analysis shows that skewed expression patterns relate to clinical outcome of COVID-19 patients. While we confirm a potentially detrimental suppression of type I IFN signalling in severe COVID-19 patients, which is linked to broad array of cell types including monocytes, NK cells, CD8^+^ T cells and plasmablasts, our results clearly suggest that regulatory events in megakaryocytic and erythroid cells may act as pivotal components of an unfavourable course of COVID-19, which mandates further prospective exploration.

## Discussion

SARS-CoV-2 infection is linked to heterogeneous clinical presentation and a variable course of the disease. While some patients suffer from severe pulmonary disease, which can ultimately lead to death due to respiratory failure and/or complications of critical illness, others may have a mild disease course or even remain asymptomatic. Our current understanding of the spectrum of the cellular responses to SARS-CoV-2 infection is still limited, particularly when it comes to a comprehension of the longitudinal cellular signatures, which are associated with the different disease trajectories, from incremental phase to convalescence. Here, we performed a longitudinal multi-omics study on PBMCs from 13 hospitalized COVID-19 patients, one additional recovery control and 14 healthy controls (cohort 1) in order to understand the dynamics of responses along a temporal axis at single-cell resolution. Obtained genomic and cellular signatures were validated in an independent single-cell RNA-seq cohort (cohort 2: 18 hospitalized COVID-19 patients and 18 healthy controls) and a separate longitudinal bulk RNA-seq cohort of 40 ICU COVID-19 patients (cohort 3: 33 survivors / 7 non-survivors). Due to the heterogeneous kinetics of the disease with regard to the onset of clinical symptoms (e.g., short vs. long incremental phase, fast vs. slower convalescence), we ordered patient samples according to a longitudinal “pseudotime”, which takes into account the WHO ordinal scale and inflammatory markers. In the cohort 1, we used an array of temporal comparison techniques (pairwise analyses between healthy and respective disease pseudotimes, longitudinal comparisons along with the categories and impulse models) and cross-validated scRNA-seq-derived signatures and bulk RNA and DNA methylation data. This longitudinal analysis provides a clear chronological rank order to some of the changes observed in prior studies: we observe a transient increase of plasmablasts and a relative decrease of memory and naïve B cells in the scRNA-seq data as well as the flow cytometry data from peripheral blood, which coincides with inflammatory severity and normalizes with convalescence. The expression of CD138 on a large fraction of plasma cells during the disease supports the theory that these cells are activated after being non-specifically mobilized from the bone marrow or other tissues. In contrast to a previous smaller study ^15^, we present evidence that plasmablasts with neutrophil-like properties are present in healthy individuals, yet in COVID-19, particularly during the critical and complicated pseudotimes, this cell population increases and contributes to the metabolic hyperactivity signal. We saw two distinct co-expression modules indicating the increased presence of plasmablasts also in the bulk data, which positively correlate with several inflammatory markers (CRP, IL-6). While the first PB module (M1) is characterized by broad GO terms relating to lymphocyte activation and cell proliferation, the second module (M2) specifically shows a compromised type I IFN signature at the peak of the disease. Downregulation of this module over time was significantly associated with a fatal outcome in cohort 3. Several interferon-related genes (*IFI6*, *IFI27*) follow a similar pattern in plasmablasts in the scRNA-seq data from cohort 1 and 2, corroborating the suppression of IFN signalling by severe SARS-CoV-2 infection in plasmablasts themselves. Of note, we found that the COVID-19 plasmablasts were predicted to be highly metabolically active in a systems biology modelling approach ^77^, beyond their expected upregulation of an unfolded protein response ^92^. Among the pathways from the transcriptional pattern-derived model, we identified glycolysis and amino acid catabolism to be transiently activated, while NAD^+^ energy metabolism, vitamin B2 and several transport processes were specifically inhibited at the critical peak of the disease in plasmablasts, when compared to other B cell subpopulations. The changes argue for the role of plasmablasts as a nutrient sink, which was already observed in extrafollicular plasmablasts as a hallmark of a systemic inflammatory response in severe malaria ^76^. Metabolic hyperactivity of plasmablasts in that study led to nutrient deprivation of the germinal center reaction, limiting the generation of memory B cell and long-lived plasma cell responses. The predicted lower energy availability at the peak of COVID-19 could indicate excessive shuttling of glucose into antibody glycosylation, which might ultimately contribute to metabolic exhaustion of the cells ^93-95^ and/or altered glycosylation patterns of antibodies, which were linked to severe COVID-19 ^96,97^.

We also found several other temporal signatures in hospitalized COVID-19 patients from the scRNA-seq data set (cohort 1), which correspond to SARS-CoV-2-induced changes observed in cross-sectional studies: we see an early and lasting depletion of NK cells and a sustained increase of the monocytic compartment, which was present until the late follow-up time point ^14,15^. We observed a lasting lymphopenia of both the CD4^+^ and CD8^+^ T cell compartment, which however was less prominently linked to the hyperinflammatory phase compared to other studies. Interestingly, we found in our DNA methylation data that hypomethylated positions were highly enriched *in cis* of canonically upregulated transcripts, which were related to positive regulation of TNF secretion, IL-1β release and innate immune signalling. *Vice versa*, downregulated transcripts comprised T cell receptor signalling and negative regulation of ATP metabolism, indicating a potential long-term regulation of the immunological misfiring ^33,98^ by epigenetic processes. Together these changes clearly suggest effects of COVID-19 immunopathology beyond the immediate cytokine release syndrome.

Most strikingly, our longitudinal approach identified two other cellular features induced by COVID-19, which were unrecognized in previous studies and are related to non-immune cell populations. First, we found a significant increase of megakaryocytes, which carried a molecular signature of a strong type I IFN response. These cells were present throughout the disease course and were associated with two longitudinal co-expression modules from the bulk RNA sequencing data (M3 and M4). Both modules were positively correlated with serum levels of D-dimers, suggesting a link between the observed megakaryocyte signature and the inflammation-induced pro-coagulative state, which has been observed as a potentially fatal complication in COVID-19 patients. Interestingly, one of the top upregulated transcripts in megakaryocytes was *IFITM3*, which encodes a protein known to confer antiviral activity in megakaryocytes and platelets ^99^. Metabolic modelling suggests that in the hyperinflammatory states, a shift towards higher rates of pyruvate metabolism and glycolysis occurs, which has been shown to sensitize platelets towards activation and aggregation ^100^. Another interesting observation was the persistent downregulation of the *TGFB1* transcript along the disease axis. Whereas *TGFB1* in platelets has been commonly associated with a pro-fibrotic function, e.g., in hypertensive cardiac remodelling ^101^, its expression in megakaryocytes orchestrates homeostatic quiescence and promotes the stress-induced expansion of haematopoietic stem cells ^86^. The observed increase of circulating HSCs and other precursor cells including megakaryocyte-erythroid progenitors (MEPs) argue for a significant influence of COVID-19 on the megakaryocytic lineage, which does not only represent a bystander response to increased platelet consumption but also contributes to viral defence and may alter aggregative properties of platelets ^31^. Although thrombopenia has been observed as a clinical correlate of critical COVID-19 (metanalysis in ^102^), it is unlikely that direct infection of megakaryocytes as seen in Dengue fever is responsible for this alteration. We found no evidence of neither *ACE2* expression in megakaryocytes nor any virus-associated reads in the cell population (data not shown). Yet, the continuous presence of type I IFN signals, as seen in our data set, has been shown to be sufficient for the exhaustion of emergency thrombopoiesis, an increase of the aggregation potential of platelets and to contribute to the vicious circle of platelet activation and wide-spread thrombus formation, e.g. in neutrophil-derived NETs ^103,104^.

The second cellular feature was primarily observed in the bulk RNA-seq data and was related to the co-expression module M7. We saw a biphasic up-regulation of a transcript group that comprises canonical components of erythropoiesis, which are most likely related to the presence of reticulocytes. GATA-1 transcription factor binding motifs linked to hypoxia-induced stress erythropoiesis ^105^ were significantly enriched in the module. We thus reasoned that this feature reflects a canonical response to hypoxia as it is present in critically ill patients and at a later stage when patients are weaned of supplemental oxygen. Post-hypoxia polycythaemia and the presence of different erythroid progenitors in the circulation has been studied as a response mechanism of the bone marrow to acute hypoxic insults and critical illness since decades ^106,107^. Mobilization of progenitor cells and their presence in the circulation has been linked to augmented immune responses ^108^. While we did not directly profile erythrocytes in the scRNA-seq data due to red blood cell lysis and size/feature filtering steps in the data processing, we found evidence for the increased presence of committed erythroid-progenitor like cells in the scRNA-seq data in two of our patients at the stage of oxygen-weaning. From the gene content and the number of cells identified, it is unlikely that these cells represent reticulocytes. Circulating bipotent megakaryocytic-erythroid progenitors (MEP) were clearly elevated along the disease trajectory and also showed clear signs of a type I IFN response. Together the results demonstrate a profound reaction of the erythroid lineage to COVID-19 at different phases of the disease.

As the last step, we examined the correlation of the potential biomarker patterns of the computed disease trajectory to clinical outcome in a larger retrospective cohort of 40 mechanically ventilated COVID-19 patients. Considering the expression change between two timepoints in patients with a comparable disease stage, we could show that two modules from cohort 1, M4 (related to megakaryocyte numbers in cohort 1) and M7 (indicative of erythropoiesis/hypoxia/platelet numbers in cohort 1), were significantly correlated with a fatal outcome in the independent cohort. Downregulation of the M2 module (associated with hypomorphic type I IFN in cohort 1) was present in both survivors and non-survivors in this cohort, corroborating the observation of failing type I interferon in severe COVID-19, but questioning its clear relation to fatal outcome. Cross-validation showed that differentially expressed genes from non-survivors were mostly contained in predefined modules from cohort 1. Whereas validated M2 markers from this cohort mapped to a broad array of immune cells in our single-cell data (monocytes, NK cells, plasmablasts), distinct clusters of validated M4 and M7 markers were expressed in megakaryocytes and committed erythroid progenitor cells in the single-cell data, respectively. The differential regulation of TFs related to megakaryocyte development and platelet production also corroborated these findings.

Overall, our longitudinal study set up provides a comprehensive view of the cellular dynamics and gene expression programs during COVID-19 using an integrated multi-omics approach in three patient cohorts. By linking changes attributed to pathophysiological events from the incremental phase of the disease to convalescence, we propose distinct events are occurring in a ranked temporal order and point to pivot points which could determine the heterogeneity of disease trajectories. Limitations of our study are given by the relatively low sample size of the initial two-centre cohort (cohort 1), which we aimed to compensate by two independent validation cohorts. The initial findings, which point to novel processes and potential biomarkers for a severe disease trajectory, mandate prospective validation.

Collectively, our study shows that circulating non-immune cells, most prominently megakaryocytes and cells of the erythroid lineage, as well as specific shifts of metabolic properties across cell types, are to be considered as an integral part of COVID-19 induced pathology. Our analysis thus provides insights into the broad effects of SARS-CoV-2 infection beyond classical immune cells and may serve as an important entry point to develop biomarkers and targeted treatments of patients with COVID-19. Our findings of interferon-activated circulating megakaryocytes may help better explaining and treating the intricate, but clinically highly relevant association of COVID-19 with thrombotic events.

## Methods

### Patients and specimen collection (cohort 1)

14 patients from two independent University hospitals (Cologne, Kiel) were recruited for the longitudinal multi-Omics study. Eligibility criteria included age ≥18 years and admission to the respective hospitals (either normal ward or ICU) with a positive SARS-CoV-2 nasopharyngeal swab by RT-PCR. Five patients were co-enrolled in an ongoing clinical trial (remdesivir) at UKSH. Enrolment for longitudinal molecular phenotyping occurred between 1st April 2020 and 6th May 2020 (seven patients at the University Medical Center Schleswig-Holstein, Campus Kiel (UKSH) and seven patients at the University Hospital Cologne (UKK), with the last sample taken on 20th May 2020). One patient with a mild disease course was recruited after his recovery to serve as an additional recovery control. The patients consented to the sampling of biomaterials, analytic processing of the biomaterials and genetic analysis, the study was approved by the independent ethical review board of Kiel University (ref no°: D 466/20) and Cologne (identifier: 20-1295). Eight healthy donors were included as controls at a single timepoint in the framework of the DZHK study (ethics vote ref no°: D 441/16). While the patients were in inpatient health care, the sampling scheme was day 1 (after admission), day 3, day 8, day 11, and day 14. At each sampling day, blood was collected in PAXgene tubes, CPT tubes, EDTA and serum monovettes (except from day 11: no CPT tubes were taken). Clinical parameters were retrieved from the electronic patients record systems or from written discharge letters from transferred patients by the COVID-19 clinical consultants (T.B., J.R.) and clinical research fellows (F.T. and P.K.).

### Validation cohorts

For validation of cellular findings from our prospective cohorts, data from two independent cohorts were analysed:

The scRNA-seq data from 18 patients admitted to the University Hospital Bonn ^14^ (cohort 2), and RNA-seq data from 40 SARS-CoV-2-positive patients admitted to the Intensive Care Unit of the Radboud university medical centre in Nijmegen (cohort 3). COVID-19 was diagnosed by a positive SARS-CoV-2 RT-PCR test in nasopharyngeal and throat swabs and/or by typical chest CT scan findings. Within cohort 3, seven patients deceased. Blood was collected in PAXgene tubes. The frozen tubes were shipped to Bonn University for mRNA sequencing. Sampling in cohort 3 was carried out in accordance with the applicable rules concerning the review of research ethics committees and informed consent in the Netherlands. All patients or legal representatives were informed about the study details and could decline to participate.

### Definition of disease severity and phases

To assess disease severity based on oxygen need and ventilation state, but also to include the inflammatory markers CRP, IL-6 and ferritin as a measure for systemic inflammation, a clinical compound scoring system was defined in accordance to the WHO Ordinal Scale and a scoring system for CRP, IL-6 and ferritin derived from observations in the LEOSS registry (see Table 2). The sum of the scores of each subcategory is used as the clinical score of the patients of each day. Based on the clinical score, disease severity phases were defined. In brief, each increase in the clinical score results in the disease phase “incremental”, while the other phases depend on the absolute clinical score (see Table 3). In case the clinical score is 9, the disease phase is set to “critical”, if the WHO Ordinal scale score is 6 or higher.

**Table 2.** Clinical Score definition. (related to Figure 1)

| **Scoring** | **Ordinal scale (WHO)** | **CRP [mg/l]** | **IL-6 [ng/l]** | **Ferritin [µg/ml]** |
| --- | --- | --- | --- | --- |
| **0** | No clinical or virological evidence of infection | < 30 | < 200 | < 1000 |
| **1** | Ambulatory, no limitation of daily activities | ≥ 30 | ≥ 200 | ≥ 1000 |
| **2** | Ambulatory, limitation of activities | ≥ 100 |  | ≥ 4000 |
| **3** | Hospitalized, no oxygen therapy | ≥ 200 |  |  |
| **4** | Hospitalized, oxygen by mask or nasal prongs |  |  |  |
| **5** | Hospitalized, non-invasive ventilation or high-flow oxygen |  |  |  |
| **6** | Hospitalized, intubation and mechanical ventilation |  |  |  |
| **7** | Hospitalized, ventilation + additional organ support – pressors, RRT, ECMO |  |  |  |
| **8** | Death |  |  |  |

Abbreviations: RRT = renal replacement therapy, ECMO: extracorporeal membrane oxygenation

**Table 3.** Pseudotime definition based on Clinical Score. (related to Figure 1)

| **Pseudotime** | **Disease Phase** | **Clinical Score** |
| --- | --- | --- |
| 0 | Uninfected (Control) | 1 |
| 1 | Incremental | Increasing |
| 2 | Critical | 9* - 13 |
| 3 | Complicated | 7 - 9* |
| 4 | Complicated | 6 |
| 5 | Moderate/early convalescence | 4 - 5 |
| 6 | Late convalescence | 3 |
| 7 | Long-term follow-up | 1 - 2 |

\* If Ordinal Scale is 6 or higher, the disease phase was classified as "critical".

### ELISA

Serum cytokines were analysed using a “Human Magnetic Luminex assay” (Biotechne, Minneapolis, Minnesota, US) with 22 analytes: APRIL, BAFF, CCL2, CCL3, CD40L, CD138, CXCL9, CXCL10, IL-1β, IL-2, IL-4 IL-6, IL-10, IL-12, IL-13, IL-17, IL-18, IL-21, IL-33, IFN-α, IFN-γ, and TNF. Frozen patient serum samples were thawed and diluted before the experiment with an equal amount of dilution buffer and the experiment was performed according to the manufacturer’s instructions. All samples were measured on a Lifematch Fluoroanalyzer (Tepnel Life Science PLC, Wythenshawe, UK) equipped with XPonent 3.1 Software (Luminex Corporation, Austin, Texas, US). Standard curves and cytokine concentrations were calculated using linear regression in Microsoft Excel GraphPad Prism (Graphpad Software Inc, San Diego, US). Serum TPO was quantified by ELISA using Human Thrombopoietin Quantikine ELISA Kit (R&D Systems, Minneapolis, Minnesota, US) according to manufacturer’s protocol.

### Anti-SARS-CoV-2 specific antibodies

Anti-SARS-CoV-2-specific IgA and IgG was quantified by CE-certified ELISA (EUROIMMUN, Lübeck, Germany).

### PAXgene™ Blood RNA Isolation and TruSeq^®^ messenger RNA (mRNA) sequencing

Blood (2.5 mL) was taken from each patient into a PAXgene™ Blood RNA Tube, containing a patented RNA stabilizer reagent composition. RNA was automated isolated in QIAGEN’s QIAcube using the PAXgene Blood miRNA Kit from QIAGEN PreAnalytiX. RNA sequencing libraries were prepared according to the Illumina TruSeq^®^ messenger (mRNA) sequencing protocol (TruSeq^®^ RNA Seq Library Prep Kit v2). The resulting libraries were sequenced on the NovaSeq 6000 (2× 50 bp, S2 chemistry).

### DNA isolation and methylation profiling

Blood (2.7 mL) was taken from each patient into an Ethylenediaminetetraacetic acid (EDTA) monovette (Sarstedt) to decelerate blood coagulation. The monovette was centrifuged for 15 min at 3000 rpm and the buffy coat was frozen in micronic tubes at −80°C. DNA was extracted using the QIAamp DNA Blood Mini Kit (Qiagen) according to manufacturer’s protocol with a QIACube (Qiagen). The Infinium© MethylationEPIC BeadChip was used to measure the DNA methylation levels. The Infinium© MethylationEPIC BeadChip targets the following regions: CpG islands, CpG sites, open chromatin, transcription factor binding sites, enhancer and miRNA promoter regions. The EPIC arrays were processed according to Illumina recommendations.

### RNA-seq data analysis

An in-house RNA-seq pipeline was used to map and align the sequenced data (https://github.com/nf-core/rnaseq). Adapters and low-quality bases from the RNA-seq reads were removed using Trim Galore (version 0.4.4), which is a wrapper tool for Cutadapt and FastQC. Reads that were shorter than 35 bp after trimming were discarded. The filtered reads were mapped to the human genome (GRCh38, gencode version 25) using STAR aligner (version 2.5.2b) ^109^. featureCounts (version 1.5.2) was used to estimate the expression counts of the genes. The expression counts were normalized across samples using the DESeq normalization method.

### Differential expression analysis

Differentially expressed genes between healthy controls and each of the COVID pseudotime samples were identified using the Bioconductor package DESeq2 (version 1.20.0). Genes with FDR adjusted *p*-value of less than 0.05, log fold change greater than 0.5 or less than - 0.5 and average expression counts of more than 100 were regarded as differentially expressed (DEGs). Longitudinal differential expression analysis of the COVID samples was performed by applying the case-only analysis from the Bioconductor package ImpulseDE2 (version 1.4.0). Pseudotimes 1, 3, 4, 5, and 6 were used as single timepoints of a time-course experiment and the patient IDs were regarded as batch effects in order to perform a paired analysis. To identify the transcripts regulated longitudinally in survivors and non-survivors from the Nijmegen cohort, differentially expressed genes between in the two selected time points were identified using DESeq2 for survivors and non-survivors separately. To perform a paired analysis, patient ID was used as batch effect.

### Co-expression analysis

Modules of co-expressed genes were identified using the WGCNA package for R (version 1.69). All differentially expressed genes identified from the pairwise and longitudinal analysis (6,318 genes in total) were used to generate the gene co-expression modules. First, pairwise gene correlations were calculated based on the log transformed normalized expression counts across all samples. A signed adjacency matrix was constructed by applying a soft threshold function with a power of 14. The Topology Overlap Matrix (TOM) constructed using the adjacency matrix was used to construct a gene tree by hierarchical clustering. Genes were then split into modules based on the gene tree by using the function cutreeDynamic with the minimum module size set to 15. Modules that were closely related were then merged using the function mergeCloseModules with parameter cutHeight set to 0.45.

To associate gene co-expression modules with clinical parameters and cell type fractions and to visualize the expression profile of the genes in a module, the module eigengene values for the samples were calculated. Spearman’s rank correlation coefficients were calculated between the module eigengenes and different clinical parameters and cell type fractions.

### Bulk BCR analyses

Bulk BCR libraries were prepared starting from 100 ng of total RNA isolated from PAXGene tubes. Library construction protocol was performed as previously described ^110^. In brief, primers for the constant (C) regions of the BCR were used during cDNA synthesis. Product was then amplified via PCR using a multiplex primer set for the variable (V) regions using the Real-Time PCR library amplification kit from KAPA Biosystems. Libraries were sequenced on Illumina MiSeq machine 2×300 bp. Sequencing reads were aligned to BCR gene reference and clonotypes were identified and grouped using the software MiXCR ^111^. Relative proportions of *IGH* classes were calculated. Alpha diversity measures were calculated using the R packages vegan and tcR (versions 1.5-6 and 2.3.2).

### Virus mRNA detection

To quantify the amount of virus present in the blood of COVID-19 patients, reads from whole blood RNA-seq data were first aligned to the human reference genome (GRCh38) using STAR with default parameters. The reads that did not map to the human genome were then aligned to the SARS-CoV-2 reference genome (NC_045512.2) using STAR with slightly relaxed parameters (--outFilterScoreMinOverLread 0.2 --outFilterMatchNminOverLread 0.2 --outFilterMatchNmin 0 --outFilterMismatchNmax 4). The reads that aligned to the SARS-CoV-2 genome with at least 40 consecutive matches were then aligned locally to the human genome using the Smith-Waterman algorithm ^112^ in order to filter any reads of human origin. The reads that aligned locally to the human genome or were composed largely of homopolymers were filtered and the remaining reads were considered as viral reads.

SARS-CoV-2-specific viral RNA from the RNA extracts from the PAXGene tubes was also quantified by RT-PCR using the RealStar® SARS-CoV-2 RT-PCR Kit RUO (altona diagnostics, Hamburg, Germany). The two amplicons were in the E gene (Primers: 5’-ACAGGTACGTTAATAGTTAA-3’, 5’-GTGTGCGTACTGCTGCAATAT-3’) and in the S glycoprotein in Spike protein 2 gene (Primers: 5’-CAGATCCATCAAAACCAAGC-3’, 5’-TCAAACAATATGGTGATTGC-3’).

### DNA methylation data analysis

DNA methylation data was analysed using the Bioconductor package RnBeads (version 1.12.1). Sites that overlapped with SNPs and had unreliable measurements were filtered resulting in the removal of 17,371 sites and 19,745 probes. 2,977 Context-specific probes, 18,976 probes on the sex chromosomes, and 4 probes with missing values were also removed. In total 41,702 out of 866,895 probes were filtered. The signal intensity values were normalized using the dasen method. Differentially methylated positions (DMPs) between healthy controls and each of the COVID-19 pseudotime samples as well as between sequential COVID-19 pseudotime samples were identified using the automatically selected rank cutoff of RnBeads.

### Functional enrichment analysis

Gene set enrichment analysis (GSEA) was conducted for the co-expression modules using GSEA desktop application. Pre-ranked analyses against Hallmark, KEGG and GO (Biological Processes) genes sets were conducted for each of the modules by ranking all genes by the module membership score. FDR of 0.05 was used as the significance threshold.

All gene ontology enrichment analyses were conducted using the Bioconductor package topGO (version 2.32.0), with all expressed genes as the universe set. In the topGO analysis, the Fisher.elim *p*-value, calculated using the weight algorithm, of 0.05 was used as the significance threshold.

Transcription factor binding sites (TFBS) enriched in the promoter regions of the co-expression module genes were identified by conducting enrichment analysis using the Bioconductor package LOLA (version 1.14.0). Promoter regions were defined as the region between 1,500 bp upstream to 500 bp downstream of the transcription start site.

Predicted transcription factor binding sites (TFBS) enriched in DMPs were identified by conducting enrichment analysis using the Bioconductor package LOLA (version 1.14.0).

### DNA methylation-transcriptome integrated analysis

For the integrated analysis of gene expression with DNA methylation, we first identified DMPs located 5,000 bp upstream and downstream of the transcription start sites of DEGs. Spearman’s rank correlation coefficient between the normalized expression count of each DEG and the methylation intensity (β-values) of its corresponding DMPs were calculated. To test the statistical significance of the correlations, we calculated the false discovery rate (FDR) using a permutation approach.

### Isolation of peripheral blood mononuclear cell (PBMC)

Blood (2 × 8 mL) was collected using venipuncture technique and processed within maximum 30 minutes. PBMCs were isolated using the BD vacutainer® cell preparation tube (CPT) with sodium citrate according to the manufacturer’s protocol. Briefly, CPT tubes were centrifuged at 1,650 × g for 20 minutes at room temperature. PBMCs were collected and washed two times with phosphate-buffered saline (PBS) and then resuspended in PBS. Half of the suspension was washed and resuspended in flow cytometry washing buffer (containing fetal bovine serum, EDTA and sodium azide in PBS) and prepared for flow cytometry within 3-5 hours. The rest of the suspension was washed once in PBS and the pellet was resuspended in resuspension medium (roswell park memorial Institute (RPMI) + 40% fetal bovine serum (FBS)), followed by freezing medium (30% DMSO in medium containing 40% FCS) according the 10× Chromium Demonstrated Protocol (Fresh Frozen Human Peripheral Blood Mononuclear Cells for Single-Cell ribonucleic acid (RNA) Sequencing, Document CG00039 Rev D). PBMCs were stored at −80°C and thawed when needed also according to the 10× Chromium Demonstrated Protocol (Document CG00039 Rev D). To prevent batch effects, all samples from different timepoints from one patient were thawed and sequenced together.

### Flow cytometry

After preparation of freshly isolated PBMCs in flow cytometry buffer, PBMCs were stained with fluorescent labelled antibodies and measured on a MACSQuant 16 flow cytometer (Miltenyi Biotec, Bergisch Gladbach, Germany). B cell subsets were stained using antibodies against CD19 (clone REA675, Miltenyi), CD20 (clone REA780, Miltenyi), CD27 (clone M-T271, Biolegend, San Diego, California, US), CD138 (clone BB4/MI15, Biolegend), HLA-DR (clone REA332, Miltenyi), IgD (clone REA740, Miltenyi), IgM (clone MHM-88, Biolegend), IgA (clone M24A, Merck Millipore), CD95 (clone DX2, Biolegend) as well as CD3 (clone OKT3, Biolegend) and CD14 (clone M5E2, Biolegend) for the dump channel. Definition of B cell populations: naïve B cells: CD19^+^CD20^+^CD27^-^; memory B cells: CD19^+^CD20^+^CD27^+^; Plasmablasts and plasma cells: CD19^+^CD20^-^CD27^hi^).

Analyses were performed using FlowJo v10 (FlowJo LLC, Beckton Dickinson, Ashland, Oregon, US) and Graphpad Prism 8 (GraphPad Software, San Diego, California USA).

### Single-cell RNA sequencing (scRNA-seq) analysis and data processing

Single-cell libraries were generated using the Chromium Next GEM Single Cell 5’ Library & Gel bead Kit v1.1 according to the manufacturer’s user guide targeting 20,000 cells per sample. The libraries were sequenced on an Illumina NovaSeq 6000 (2×100 bp, S4 chemistry) to generate > 500 million reads per library. Additionally, the Chromium Single Cell V(D)J Enrichment Kit for human B cells were applied together with the Chromium Single Cell 5’ Library Construction Kit. Those resulting libraries were sequenced on an Illumina NovaSeq 6000 (2× 150 bp, S4 chemistry) to generate > 50 million reads per library. The sequences were processed using cell ranger v3.1.0 (10× Genomics). Each sample was mapped to GRCh38 *Homo sapiens* reference genome, in order to produce their respective count matrices.

### scRNA-seq data quality control and data analysis

Raw feature-barcode matrixes were filtered using Seurat package (version 3.1.5) in R environment ^113,114^; low quality cells that were potentially disrupted or doublets cells were removed from the analysis using number of features (number of reads mapping to gene between [200;5000]) or percentage of mitochondria (lower than 25%).

Each filtered sample matrix was then merged into a single object containing 358,930 cells with overall reads mapping to 22,519 human genes. The merge object was normalized and scaled using LogNormalized() and ScaleData() functions respectively. Principal component analysis was performed utilizing the top 2,000 variable genes. We identified the clusters using the standard *k*-nearest neighbour method based on 80 dimensions with a 0.2 resolution. In total 37 clusters were displayed as a Uniform Manifold Approximation and Projection (UMAP).

### scRNA-seq signature genes

Cluster cell types were identified by their corresponding gene signatures using the Wilcoxon rank sum, with a cut-off based on genes expressed in more than 25% of the cluster cells and exhibiting a 0.25-fold difference between clusters. Clusters of interest were identified based on marker genes; monocytes (*CD14*), granulocytes (*FCGR3B*), erythroid cells (*HBA1* and *HBA2*), NK cells (*GNLY*), proliferative lymphocytes (*MKI67*), CD4^+^ T cells (*CD4*), CD8^+^ T cells (*CD8A*), dendritic cells (*PLD4*), B cells (*CD19*), plasmablasts (*CD38* and *MZB1*), megakaryocytes (*ITGA2B* and *TUBB1*) and cell precursors (*CD34*), such as, haematopoietic stem cells (HSCs), megakaryocyte-erythroid progenitor cells (MEPs), common myeloid progenitors (CMPs) and granulocytes-macrophage progenitors (GMPs). To confirm our findings, we used SingleR (version 1.0.6) R package that assigns each individual cell to a known cell type based on transcriptome reference datasets (NovershternHematopoieticData and BlueprintEncodeData reference datasets) ^36^. Cell type proportions were quantified per sample (Supplementary Table 2) and grouped based on pseudotimes. We compared cell type proportions of healthy controls against patients using a Mann-Whitney non-parametric test and measured cell proportion changes between pseudotimes by comparing a linearlinear mixed model with pseudotime (proportion of cell type~pseudotime +[1|patientID]) and compared it against a reduce model without pseudotime (proportion of cell type +[1|patientID]) by the means of an ANOVA (Figure 2f). We found most cell types to be impacted by disease trajectory, with only proliferative lymphocytes and cell precursors not having a significant difference between pseudotimes. Furthermore, we correlated cell type proportion with the clinical parameters available for each sample using spearman correlation (Figure 2g).

### Cell type specific analysis

The clusters identified as cell types of interest-B cell compartment, megakaryocytes and cell precursors were pulled from the merged object. Each cell type of interest was re-clustered and analysed separately. B cell compartment and cell precursors were re-clustered using 80 PCs, while megakaryocytes and their respective precursors (HSCs and MEPs) were re-cluster using 60 PCs. B cell compartment clusters were assigned based on expression of marker genes, with memory B cell expressing *CD73/NT5E*, naïve B cells expressing *IGHD* and *CD185/CXCR5*, transitional B cells expressing *CD9*, plasmablasts expressing *CD27* and *CD38*) and neutrophil-like cell expressing *ELANE, MPO* and *CAMP* ^73^. We presented the B cell compartment as a cell trajectory analysis using monocle3 (Qiu et al., 2017). Trajectories were calculated and the cells displayed based on monocle3 pseudotime approach rooted on the previously identified transitional B cells. Plasmablasts (with neutrophil-like cells included), megakaryocytes and cell precursors signature genes for the individual disease groups were selected based on genes expressed in more than 25% and 0.25-fold difference between pseudotimes. Differentially expressed genes between healthy controls and each of the COVID pseudotimes was identified using MAST ^115^ and GO enrichment analysis was performed with TopGO package for R (version 2.38.1) ^116^ and GO terms of interest selected based on fisher classic test statistic (*p*-value < 0.05).

To validate our findings, we performed a parallel analysis from an independent cohort (cohort 2) of mild and severe COVID-19 patients using another scRNA-seq technology (Rhapsody™ BD) (Schulte-Schrepping et al., 2020). Similarly, we identified plasmablasts and megakaryocytes, identified differences in cell proportions and tested if genes of interest were expressed differently based on their disease classification (control, mild COVID-19 or severe COVID-19 patients).

### single cell BCRseq (scBCR-seq) analysis

BCR sequences were processed using cellranger v3.0.1 vdj function. Relative proportions of *IGH* classes were calculated. Alpha diversity measures were calculated using the R packages vegan and tcR (versions 1.5-6 and 2.3.2). scBCR-seq information was merged with B cell compartment containing both B cells and plasmablasts using barcode information. BCR information was merged with scRNA-seq expression, thus, we were able to discern IG class information per cell type and displayed it in the form of a UMAP or monocle3 cell trajectory.

### Metabolic modelling

Blood metabolites originating from the Human Metabolome Database (HMDB)^117^; specifically, an advanced search of blood metabolites from healthy adults was initially conducted in March 2019. In the same time period, a list of all metabolites in Recon 3D ^118^ was downloaded from Virtual Metabolic Human (VMH) database ^119^. The latter contains the HMDB indices of the compounds, which enabled the merging of the two databases into one (Spring 2020). Compounds were removed from the database (e.g., drug-related ones or without FooDB IDs (https://foodb.ca/)) and the metabolites nomenclature was altered to be compatible with an adapted version of the model Recon 2.2 ^120^. All values were transformed to mM to correspond with mean values of the HMDB database (cut-off of values > 10^-6^). Water was set to 55,000 mM, and pH was set to 7.4. All calculations were conducted with the R packages sybil (version 2.1.5) ^121^ and sybilSBML (version 3.0.5) ^121^ along with their dependencies.

### Reconstruction of tissue-specific metabolic models from bulk sequencing data

For the reconstruction of metabolic models from bulk sequencing data we used a previously described two-step approach that first discretizes gene expression based on differential gene expression analysis and subsequently reconstructs metabolic models based on gene expression states ^53,122^. Differentially expressed genes between healthy controls and each of the COVID pseudotimes from previous differential expression analysis were used separately to build generalized linear models and tested with disease state as an independent variable. Wherever possible, i.e. when models reached full rank, we included the donor identifier to control for paired samples with similar genetic background in the data. The significance cutoff used for optimizing independent filtering was adjusted to α = 0.05 before differentially expressed genes were extracted from the tests. For each gene, based on the directionality of changes in gene expression activity between conditions and the significance of changes, we determined a binary activity (*on* or *off*) if a gene had at least one case of significant change in activity (adjusted *p*-value < 0.05) between any pair of conditions.

Subsequently, we used the binary gene activity as an input into the iMAT approach ^123^ on a generic metabolic model of humans (Recon 2.2)^120^ constrained with the serum metabolic environment. In order to test method-inherent uncertainties in the reconstructed context-specific metabolic network, the model reconstruction procedure was repeated fifty times while leaving out gene activity data for 5% of the genes each time.

### Reconstruction of cell-specific metabolic models from single-cell sequencing data

Reconstructions of cell-specific metabolic models were created by integrating single-cell transcriptomics with a human genome-scale metabolic network ^120^ conditioned with the serum metabolic environment. We employed a two-step approach, in which *StanDep* ^77^ first identifies a core reaction list across cell types. Second, the FASTCORE algorithm ^124^ in the COBRA Toolbox v.3.0 ^125^ then builds context-specific models defined by sets of active core reactions in the extracted model. scRNA counts from patient, megakaryocytes, plasmablasts, memory B cells and naïve B cells were used as input for StanDep. Normalized counts were converted into TPM-values, and ENSEMBL gene names were mapped to Recon 2.2 ^120,126^. For StanDep, expression data from identified core genes across cell types were used to calculate enzyme type and expression within the model. Enzyme expressions were log10 transformed and counted as a binary matrix (rows representing enzymes and columns as bins) to identify the minimum and maximum enzyme expression values. A complete linkage metric for hierarchical clustering with Euclidean distance was used to cluster (number of clusters = 40) genes based on gene expression. Assembled core reaction matrices were defined and input into FASTCORE to reconstruct context-specific metabolic models. Based on an updated version of Recon 2.2 simulated in the serum metabolic environment, a consistent model was generated using FASTCORE’s FASTcc algorithm in MATLAB. With this consistent model, the assembled core reaction matrices, and additional optional core reactions, such as the biomass objective function from Recon 2.2, an input file was generated for every cell-specific model comprising at least 30 core reactions. FASTCORE processed these input files, together with the consistent Recon 2.2 model, and the default value 10^-4^ for ε to generate a list of all required reactions for each cell-specific metabolic model. With these lists, new metabolic models were curated and optimized for the biomass objective function and/or the viral biomass objective function.

### Identification of disease-specific metabolic pathways

Tissue-and cell-specific models were stratified by cell/tissue type (megakaryocytes, plasmablasts, memory B cells, naïve B cells and NK cells) and annotated with clinical metadata (COVID-19 disease status) according to donor and sampling timepoints. Reactions per pathway and cell were counted for a total of 82 metabolic pathways that were identified in the models. For each of the four model types, pathways were filtered out if reaction counts were zero across all models of that type. Differential pathway activity was determined for each pathway by comparing reaction counts across all eight pseudotimes via Kruskal-Wallis test as implemented in the R-package coin (parameters: two-sided test, unpaired, average-scores for ties, and without continuity correction). Resulting probability values were corrected for multiple testing via the Benjamini and Hochberg method. Significantly differential active pathways across disease states were determined via this method for all four model types (total PBMC, B cells, megakaryocytes, and plasmablasts) separately with an FDR cut-off of ≤ 0.05. For the heatmap representation, pathways that were identified as significantly differential active in at least two of the four model types were selected and ordered by *p*-values of the total PBMC models.

### Transcription factor activity analysis

Putative transcription factor activity from RNA-seq data was assessed per pseudotime against healthy controls using the human gene set resource DoRothEA v1, which provides a curated collection of transcription factor and target genes interactions (the regulon) from different sources ^127^. Only interactions with high, likely, and medium confidence (levels A, B, C) were considered. Regulons were statistically evaluated using the R package viper (v1.22.0; row-wise *t*-tests) and regulons having at least 15 expressed gene targets were considered ^128^. Identification of upstream regulatory signalling pathways from downstream gene expression was performed on *t*-statistic values from viper against the Omnipath interaction database (R package OmnipathR version 1.2.1) applying CARNIVAL (version 1.0.1 with IBM Cplex solver as network optimizer) ^47^. For the resulting network, only edges with an inferred weight > 50 (on a scale from 1 - 100) were considered. The similarity between the transcription factor activity of cohort 1 and cohort 3 was accessed using Pearson correlation. The significance of the correlation was confirmed by a bootstrapping approach. We permuted gene names row wise 1,000 times and correlated each time the inferred TF activity between the original and permuted gene expression. The resulting distribution of correlation values was fitted to a skewed gaussian normal distribution, finding a standard deviation of 0.08.The significance in the differential regulation of transcription factors over time in the non-survivor vs. the survivor groups of cohort 3 was quantified via a moderated t-test using limma ^91^, while accounting for patient correlation using a block design and limma’s duplicateCorrelation function.

## Data Availability

As of now, data will be available upon request.

## Acknowledgements

We thank Melanie Schlapkohl, Catharina von der Lancken and Melanie Vollstedt for perfect technical assistance. This work was supported by the German Research Foundation (DFG) CCGA Nr. 07495230, ExC 2167 Precision Medicine in Chronic Inflammation (RTFVI), the research group and the CRC1182 C2 project to P.R., INST 37/1049-1,INST 216/981-1, INST 257/605-1, INST 269/768-1, INST 217/988-1, INST 217/577-1, and EXC2151/1 to J.L.S.; SFB TR57 and SPP1937 to J.N.; Helmholtz-Gemeinschaft Deutscher Forschungszentren, Germany (sparse2big to J.L.S.), EU projects SYSCID (733100 to P.R. and J.L.S.), the DZIF, Germany (TTU 04.816 and 04.817 to J.N.); the Hector Foundation (M89 to J.N.). AD acknowledges support by the DFG through the Cluster of Excellence Controlling Microbes to Fight Infections (CMFI, EXC 2124) and the German Center for Infection Research (DZIF). CK acknowledges support by the DFG through EXC 2167, sub-project RTF-VIII and the CRC 1182.

We are indebted to the patients, their families and the hospital staff for support, without whom this study would not have been possible.

## Consortia

Deutsche COVID-19 Omics Initiative (DeCOI)

Angel Angelov, Robert Bals, Alexander Bartholomäus, Anke Becker, Daniela Bezdan, Ezio Bonifacio, Peer Bork, Thomas Clavel, Maria Colome-Tatche, Andreas Diefenbach, Alexander Dilthey, Nicole Fischer, Konrad Förstner, Julia-Stefanie Frick, Julien Gagneur, Alexander Goesmann, Torsten Hain, Michael Hummel, Stefan Janssen, Jörn Kalinowski, René Kallies, Birte Kehr, Andreas Keller, Sarah Kim-Hellmuth, Christoph Klein, Oliver Kohlbacher, Jan O. Korbel, Ingo Kurth, Markus Landthaler, Yang Li, Kerstin Ludwig, Oliwia Makarewicz, Manja Marz, Alice McHardy, Christian Mertes, Markus Nöthen, Peter Nürnberg, Uwe Ohler, Stephan Ossowski, Jörg Overmann, Silke Peter, Klaus Pfeffer, Anna R. Poetsch, Alfred Pühler, Nikolaus Rajewsky, Markus Ralser, Olaf Rieß, Stephan Ripke, Ulisses Nunes da Rocha, Philip Rosenstiel, Antoine-Emmanuel Saliba, Leif Erik Sander, Birgit Sawitzki, Philipp Schiffer, Eva-Christina Schulte, Joachim L. Schultze, Alexander Sczyrba, Oliver Stegle, Jens Stoye, Fabian Theis, Janne Vehreschild, Jörg Vogel, Max von Kleist, Andreas Walker, Jörn Walter, Dagmar Wieczorek, John Ziebuhr

## Author Contributions

J.P.B., N.M. and F.T. contributed equally and are listed in alphabetical order. T.B., L.B., J.I.B., D.B., J.F., U.G., J.J.S., P.K., A.K. and E.R. contributed equally and are listed in alphabetical order. H.B., B.H., C.K., J.S., J.H., M.K., J.R., S.S. and P.R. share senior authorship. S.S., J.S. and P.R. conceived study concept and design. J.P.B., N.M., F.T., T.B., U.G., P.B., G.E., A.F., N.F., R.J., K.F.R, J.R., A.S., H.B., B.H., C.K., J.H., M.K., J.R. contributed to study concept and design. J.P.B., N.M., F.T., L.B., U.G., A.K., E.R., J.J.S., H.B., B.H., C.K. and P.R. contributed to literature search, data interpretation and writing the initial manuscript. All authors contributed to reviewing and editing of the manuscript. F.T., T.B., J.I.B., J.F., U.G., P.K., E.R., J.D., G.E., J.F., S.F., N.F., J.F., A.G., J.H., F.H., S.I., S.K., C.L., G.L., M.L., R.M., J.N., P.P., C.R., J.R., A.S., D.S., D.S., J.H., M.K. and J.R. participated in sample collection and processing. J.P.B, N.M., F.T., L.B., U.G., J.J.S., A.K., E.R., A.C.A., N.B., T.B., B.B., A.D., D.E., M.F., M.P.H., Y.H.K, R.K., C.L., G.M., A.R., J.S.S., T.U., K.P.W., M.W., J.Z., H.B. and C.K. participated in data analysis. J.P.B, N.M., F.T., L.B., D.B., U.G, J.J.S, A.K., E.R., H.B. and C.K. made figures and tables.

## Declaration of interests

The authors do not declare any conflict of interest.

**Supplementary Figure 1 related to Figure 2**

**a**, SingleR UMAP representation of all merged samples. In total, 358,930 cells are depicted. Cells are coloured by Single R qualification.

**b**, Cluster UMAP representation of all merged samples. In total, 358,930 cells are depicted. Cells are coloured by cluster.

**c**, Expression of marker genes of interest in UMAPs of all merged samples. *CD14* expression (top left), *CD3E* expression (top centre), *CD8A* expression (top right), *CD4* expression (bottom left)*, CD19* expression (bottom centre) and *MKI67* expression (bottom right) are depicted. *CD14* marker for monocytes, *CD3E* marker for T cells, *CD8A* marker for CD8^+^ T cells, *CD4* marker for CD4^+^ T cells, *CD19* marker for B cells and *MKI67* for proliferative cells.

**Supplementary Figure 2. related to Figure 3 and Figure 4**

**a**, Number of significantly differentially expressed genes (DEGs) between controls and COVID-19 pseudotimes and number of longitudinal DEGs across different pseudotimes. Colours discriminate upregulated (red) and downregulated (blue) genes in COVID-19 samples compared to controls and longitudinal DEGs (purple).

**b**, Heatmap showing genes differentially expressed between critical (pseudotime 2) and complicated (pseudotime 3) disease. The normalized gene counts are scaled by row and row-wise *z*-scores are plotted in the heatmap.

**c**, Heatmap depicting the activity of top 50% most variable transcriptions factors (TFs) across the seven pseudotimes relative to the controls. A star within a cell denotes a significant (*p*-value < 0.05) activation or inhibition of the respective TF.

**d**, Inference of upstream signalling networks. Left: directed nearest neighbour network based on inferred hub-genes MAPK1/3 for pseudotime 3. The node size corresponds to the node degree. Violet and red node colors correspond to active/inactive nodes, respectively. Edge colors black and red correspond to activating and inhibiting interactions. Right: Heatmap depicting the degree centrality of all inferred networks across the pseudotimes. Rows have been clustered by complete linkage according to their Eucledian distance.

**e**, Disease-state specific total metabolic activity for bulk RNA-seq data showing the total number of active reactions for context-specific metabolic models.

**f**, Eigengene heatmap of co-expression modules constructed using all pairwise and longitudinal differentially expressed genes. Columns are ordered by pseudotimes and annotated by age, gender and patient IDs.

**Supplementary Figure 3. related to Figure 5**

**a**, RnBeads computational deconvolution of whole blood samples from COVID-19 patients and controls based on EPIC array data.

**b**, Comparison of mean methylation intensities (β-values) between pseudotimes 3 and 4, 4 and 5, and 5 and 6. Significant DMPs, defined by the automatically selected rank cut-off by RnBeads, are highlighted in red (hypermethylated) and blue (hypomethylated).

**c**, Number of significant DMPs between subsequent COVID-19 pseudotimes. Colours discriminate hypermethylated (red) and hypomethylated (blue) sites in later COVID-19 pseudotimes compared to former.

**d**, Heatmap showing the significant enrichment, quantified by odds ratio, of transcription factor binding sites (TFBS) in the DMPs identified between pseudotimes 3 and 4, 4 and 5, and 5 and 6. Order of the heatmaps correspond to the comparison depicted in panel **b**. Selected top transcription factors are visualized.

**e**, Histogram depicting the direction of correlation between DMP-DEG pairs. DMP-DEG correlations with FDR < 0.05 are visualized.

**f**, Heatmap showing the average expression of canonical DMP-DEGs upregulated in active disease (pseudotimes 1, 2, 3 and 4) in different cell-types of cohort 1 (from scRNA-seq data). The average expression in severe stages of the disease (pseudotimes 1, 2 and 3) is shown. The average genes counts are scaled by row and row-wise z-scores are plotted in the heatmap. Genes are labelled by the co-expression modules and are hierarchically clustered for each module separately.

**g**, Heatmap showing the average expression of canonical DMP-DEGs downregulated in disease recovery (pseudotimes 5 and 6) in different cell-types of cohort 1 (from scRNA-seq data). The average expression in recovery stages of the disease (pseudotimes 5 and 6) is shown. The average genes counts are scaled by row and row-wise z-scores are plotted in the heatmap. Genes are labelled by the co-expression modules and are hierarchically clustered for each module separately.

**Supplementary Figure 4. related to Figure 6**

**a**, Cell trajectory analysis of B cell compartment. Cell trajectory was calculated using Monocle3 and the cells coloured by IG class.

**b**, Cell trajectory analysis of B cell compartment. Cell trajectory was calculated using Monocle3 and the cells coloured by pseudotime.

**c**, Flow cytometry analysis of Plasmablasts. Plasmablasts were stained for HLA-DR and CD138. Double positive HLA-DR^+^/CD138^+^ cells on top panel, CD138^+^ cells on center panel and HLA-DR^+^ cells on bottom panel. Proportions of plasmablasts were plotted against the sampling time relative to the disease onset. The points were coloured by corresponding pseudotime and connected by patient. Only patients from Kiel cohort (*n* = 7 individuals) are depicted.

**d**, BCR diversity measures classified by pseudotime. Data based on bulk BCR-seq analysis. BCR diversity as number of clones (top left), inverse Simpson index (bottom left), Gini inequality index (top right) and clonality (bottom right).

**e**, BCR diversity measures for B cell subtypes classified by pseudotime. Data based on scBCR-seq. Diversity measures grouped by B cell subtypes memory B cells and plasmablasts. BCR diversity as number of clones (top left), inverse Simpson index (bottom left), Gini-Simpson index (top right) and clonality (bottom right).

**f**, IGHA BCR proportion classified by pseudotime. Data based on bulk BCR-seq analysis.

**g**, Gene expression of genes related to IGHA by pseudotimes. *IGHA1* and *IGHA2* expression classified by pseudotimes (colour) for Memory B cells (top) or plasmablast (bottom).

**h**, IGHG BCR proportion classified by pseudotime. Data based on bulk BCR-seq analysis.

**i**, Gene expression of genes related to IGHG by pseudotimes. *IGHG1* and *IGHG2* expression classified by pseudotimes (colour) for Memory B cells (top) or plasmablast (bottom).

**j**, IGHA BCR proportion for B cell subtypes. Memory B cells on top and plasmablast on bottom classified by pseudotime. Data based on scBCR-seq analysis.

**k**, Heatmap for proportion of V usage of IGHA. Top genes related to IGHA1 or IGHA2 by pseudotime. Usage frequency depicted by colour intensity. Data based on bulk BCR-seq analysis.

**l**, IGHV3-23 expression in plasmablasts during disease trajectory. For each UMAP, IGHV3-23-expressing cells were highlighted (red).

**m**, IGHG BCR proportion for B cell subtypes. Memory B cells on top and plasmablast in bottom classified by pseudotime.

**n**, Heatmap for proportion of V usage of IGHG. Top genes related to IGHG1 or IGHG2 by pseudotime. Usage frequency depicted by colour intensity. Data based on bulk BCR analysis.

**o**, IGHV3-30 expression in plasmablasts during disease trajectory. For each UMAP, IGHV3-30-expressing cells were highlighted (dark red).

**Supplementary Figure 5. related to Figure 7**

**a**, Schematic workflow of the scRNA-seq analyses performed on the cell precursors identified in Figure 2.

**b**, Cell precursors represented as a UMAP. In total 779 cells are depicted. Clusters represented are HSCs (pink), MEPs (yellow), CMPs (dark yellow) and GMPs (light orange).

**c**, Cell precursors pseudotimes represented as a UMAP. In total 779 cells are depicted.

**d**, HSCs-specific proportions grouped by disease phases. Points represent individual samples and horizontal bars the mean of a particular pseudotime. Pseudotimes are represented by colours.

**e**, MEPs-specific proportions by disease phases, pseudotimes are represented by colours.

**f**, GMPs-specific proportions grouped by disease phases. Points represent individual samples and horizontal bars the mean of a particular pseudotime. Pseudotimes are represented by colours.

**g**, CMPs-specific proportions grouped by disease phases. Points represent individual samples and horizontal bars the mean of a particular pseudotime. Pseudotimes are represented by colours.

**h**, Dot plot for signature genes in cell precursors. Genes were selected based on ten most characteristic upregulated genes. Colour discriminates upregulated (red) or downregulated (blue) genes, while point sizes represents the number of cells per group expressing the corresponding gene.

**i**, Gene expression of genes of interest along the disease axis. Violin plot based on pseudotime expressions classified by colour for *BST2*, *IFITM1*, *IFITM2* and *IFI6*.

**j**, HSCs and MEPs proportions grouped by disease state. Points depicting samples classified as healthy controls in green, as inflammatory stages (pseudotimes 1-3) in blue or as convalescent stages (pseudotimes 4-7) in red. Statistics based on Mann-Whitney U test.

**Supplementary Figure 6. related to Figure 7**

**a**, Number of active reactions during disease course. Number of reactions calculated based on the metabolic model for plasmablasts (red), memory B cells (blue), naïve B cells (green) and megakaryocytes (purple). Number of active reactions corrected for cell number.

**b**, Overview of fold-changes in metabolic activity of individual reactions around spermidine degradation pathway in megakaryocytes. Red indicates reactions that are more frequent in megakaryocytes from severe time points vs. the healthy time point, blue indicates no change or down-regulation.

**c**, Overview of fold-changes in metabolic activity of individual reactions around pyruvate and methyglyoxal metabolism pathway in megakaryocytes. Red indicates reactions that are more frequent in megakaryocytes from severe time points vs. the healthy time point, blue indicates no change or down-regulation.

